# Clinical characterization of human monkeypox infections in the Democratic Republic of the Congo

**DOI:** 10.1101/2022.05.26.22273379

**Authors:** Phillip R. Pittman, James W. Martin, Placide Mbala Kingebeni, Jean-Jacques Muyembe Tamfum, Qingwen Wan, Mary G. Reynolds, Xiaofei Quinn, Sarah Norris, Michael B. Townsend, Panayampalli S. Satheshkumar, Bryony Soltis, Anna Honko, Fernando B. Güereña, Lawrence Korman, John W. Huggins, The Kole Human Monkeypox Infection Study Group

## Abstract

We describe the results of a prospective observational study of the clinical natural history of human monkeypox virus (MPXV) infections at the remote General Reference Hospital of Kole (Kole hospital), the rainforest of the Congo River basin of the Democratic Republic of the Congo (DRC) from March 2007 until August 2011. The research was conducted jointly by the Institute National de Recherche Biomedical (INRB) and the US Army Medical Research Institute of Infectious Diseases (USAMRIID). The Kole hospital was one of the two previous WHO Monkeypox (MPX) study sites (1981-1986). The hospital is staffed by a Spanish Order of Catholic Nuns from La Congregation Des Seours Missionnaires Du Christ Jesus including two Spanish physicians, who were members of the Order as well, were part of the WHO study on human monkeypox. Of 244 patients admitted with a clinical diagnosis of MPXV infection, 216 were positive in both the Pan-Orthopox and MPXV specific PCR. The cardinal observations of these 216 patients are summarized in this report. There were three deaths (3/216) among these hospitalized patients; fetal death occurred in 4 of 5 (80%) patients who were pregnant at admission. The most common complaints were rash (96.8%), malaise (85.2%), sore throat (78.2%), and lymphadenopathy/adenopathy (57.4%). The most common physical exam findings were MPX rash (99.5%) and lymphadenopathy (98.6%). Age group of less than 5 years had the highest lesion count. Primary household cases tended to have higher lesion counts than secondary or later same household cases. Of the 216 patients, 200 were tested for IgM & IgG antibodies (Abs) to Orthopoxviruses. All 200 patients had anti-orthopoxvirus IgG Abs; whereas 189/200 were positive for IgM. Patients with hypoalbuminemia had a high risk of severe disease. Patients with fatal disease had significantly higher maximum geometric mean values than survivors for the following variables, respectively: viral DNA in blood (DNAemia, p=0.0072); maximum lesion count (p=0.0025); day of admission mean AST and ALT (p=0.0002 and p = 0.0224, respectively, adjusted p-values).

**Author Summary:** This is a prospective observational study of Human monkeypox disease, an emerging infectious disease in parts of the continent of Africa. There are certain differential characteristics when compared to other pox diseases. This paper describes the presenting symptoms and signs of human monkeypox disease, laboratory findings and makes recommendation for the medical treatment of patients with monkeypox disease.

## Introduction

Monkeypox virus (MPXV), a zoonotic orthopoxvirus (OPXV), causes a potentially lethal infection in humans that clinically resembles smallpox (1). Since the eradication of smallpox in the 1970’s (2), MPXV has been considered the OPXV posing the greatest danger to human populations (3). Smallpox vaccination provides partial cross-protective immunity against human monkeypox (MPX) (4–6). The cessation of routine vaccination against smallpox has left human populations increasingly vulnerable to MPXV, and to variola virus (VARV, causative agent of smallpox), a potential agent of biowarfare (7).

Historically, most cases of MPX have occurred in western and central Africa, primarily the Democratic Republic of the Congo (DRC, formerly Zaïre) (3, 8, 9). Intensive surveillance for human MPX from 1981 to 1986 (4) and stochastic modeling of MPXV transmission in human populations concluded that human to human transmission of monkeypox did not constitute a major public health problem. However, more recent active surveillance undertaken in 2005– 2007 demonstrated an average annual cumulative incidence of 5.53 per 10,000, a 20-fold increase over 30 years (11). This finding has raised concerns over the potentially growing threat posed by MPXV and other OPXVs in the context of an increasingly immunologically naïve human population (12). The re-emergence of human monkeypox infection, from 2017-2019, in West and Central Africa supports this proposition (13–16). Furthermore, cases of monkeypox disease in humans have been imported into England, Singapore and Israel (17–19), in addition to the human cases in the United States stemming from rodents imported from Ghana (20) and a traveler from Nigeria to Dallas, TX (21). More recent human-to-human transmission modeling of monkeypox by Grant, et al., concluded, “The geographic spread of monkeypox cases has expanded beyond the forests of central Africa, where cases were initially found, to other parts of the world, where cases have been imported. This transmission pattern is likely due to the worldwide decline in orthopoxvirus immunity, following cessation of smallpox vaccination.” They further used mathematical modeling to detail that “Monkeypox could therefore emerge as the most important orthopoxvirus infection in humans… [and] the epidemic potential of monkeypox will continue increasing” (22).

The original intent of the study was to obtain human MPX infection data to include such parameters as lesion counts, levels of viremia and basic clinical lab tests to compare human MPX infection to various orthopox infection animal models of human smallpox. Since this study was initiated researchers have developed an approved treatment for variola; however, the continued use of such animal models will be important in exploration of additional therapeutic options for smallpox. The biodefense community recognizes a need for continued therapeutics development in the event of VARV reintroduction to the world as a human infection.

A greater understanding of human MPXV infection will aid efforts to protect human populations from the threat posed by MPXV as well as the potential threat posed by VARV (23). In this study, we sought to improve understanding of the clinical course of human MPXV infection which will be crucial for the continued development of therapeutic interventions against human OPXV infections.

## Methods

### Study site and population

This study site was Kole hospital in the Sankuru District of Kasaï-Oriental Province in DRC. Land cover in the Sankuru District consists of tropical rainforest, savannah, and traditional agricultural fields. Residents of the district are primarily subsistence farmers and hunters who live in small villages that average 100 individuals, spread amongst small clearings in the forest and small farming communes of extended families of less than 15 people (11). The research was conducted jointly by the Institut National de Recherche Biomédicale (INRB) and the US Army Medical Research Institute of Infectious Diseases (USAMRIID) at one of the two previous WHO MPX study sites (1981-1986).

Admission to the “Monkeypox Isolation Ward” was based upon clinical diagnosis of human MPXV infection by hospital staff. A clinically overt case of active MPX was defined as having either (a) vesicular rash, with crops of vesicles of similar developmental stage in each body region (regional monomorphism) typically first appearing on the face, hands, and feet with centrifugal distribution (pox-like rash) or (b) fever of up to 39°C or a history of fever, rash, lymphadenopathy, headache, malaise, or prostration in the past 2 weeks for which there is no attribution. These patients were informed about the observational study and given the option to participate by granting informed consent or if a minor, parental permission or ascent as the situation required.

Patients were typically accompanied by family members, who provided basic care for the patient. During this study family members stayed with the admitted patients within the isolation compound where they prepared food for patients and family members.

### Compliance statement

The study protocol was reviewed and approved by the Human Use Committee at the USAMRIID (FY05-13) and the Headquarters, United States Army Medical Research and Development Command Institutional Review Board (IRB) as well as the Ethics Committee at the University of Kinshasa School of Public Health (KSPH). The study was conducted according to the approved protocol and applicable U.S.A. federal, DOD and local regulatory requirements and guidelines as well as in compliance with applicable Congolese law. The study was conducted under the oversight of the Ministry of Health with appropriate guidance and collaboration from the KSPH and the INRB. All personnel involved in the study had human subjects protection training. Patient’s privacy was respected in keeping with local cultural and hospital standards. Reasonable care was taken to safeguard subject confidentiality and protect medical information consistent with limitations of the hospital facility. Informed consent/assent was obtained before any study procedure was performed. For children aged 0-11 years, written informed consent was obtained from parent or guardian. For minors 12-17 years, written assent was obtained as well as written parental/guardian informed consent. All were given a copy of their signed informed consent/assent.

### Study design

Patients who presented to hospital admitting with a presumptive diagnosis of MPX were admitted to the physically separate infectious disease ward and offered an opportunity to be evaluated for enrollment in the study but received the same treatment independent of enrollment.

Once enrolled in this prospective observational study, continuation was based upon obtaining positive results with Pan-OPXV PCR. All Pan-OPXV PCR positive patients were also positive in the MPXV specific PCR. Positivity in any tissue was considered sufficient for inclusion in the PCR positive group.

Once consent had been obtained, study personnel obtained a medical history; performed a physical examination; recorded vital signs and weight; conducted a complete lesion count; and collected specimens (blood, throat swabs, lesion (pox) swabs, and urine) (scabs were collected when brought in by patient) for viral load PCR analysis, viral culture, and hematology at enrollment and several time points throughout hospitalization: up to day 14, on the day of discharge, and approximately day 75. A pregnancy test was administered on day 0 to female participants of childbearing age but pregnant subjects were still allowed to enroll and participate. Onsite physicians examined patients on day of admission and the following 3 days, as well as the day of discharge and upon follow-up at day 75 (±15 days). The duration of hospitalization varied but was typically between 7 and 21 days. The severity of a given symptom or sign (mild, moderate, severe) upon admission or during hospital stay was not collected. As a substitute for this design flaw, we used number of symptoms and signs as a “surrogate” of severity and labeled as level 1, level 2, level 3 or level 4 (death).

### Adjunctive medications administered to patients under direction of hospital staff

In recognition that many patients were hospitalized with fever which may include other infectious processes in addition to or in lieu of MPX, patients were empirically treated with amoxicillin for the first few days until the etiology of the fever was clearly established. If the patients, had clinical suspicion of pneumonia or sepsis, the patients were treated with other available antibiotics. Because the Congo River basin has very high endemicity for Falciparum malaria, all patients with fever were treated empirically with appropriate antimalarial agents until such time as sequential malaria smears confirmed there was no detectable parasitemia or a full course of malaria treatment was completed if parasitemia was confirmed. A 10% solution of Potassium Permanganate was used to disinfect skin in conjunction with bathing and other hygienic measures to mitigate contact and fomite transmission of the infection.

Aspirin, acetaminophen and diclofenac were routinely given to reduce fever and treat pain, including painful adenopathy which patients frequently experienced. Mebendazole was given empirically on admission and continued for those who screened positive for evidence of helminth infection.

One of the most significant interventions that the hospital staff provided was nutritional support, which was given not only to the hospitalized patients, but also to those accompanying family members who provided primary nursing care, hygienic measures and emotional support to the hospitalized patients. When routine dietary intake was compromised by oropharyngeal lesions and painful cervical adenopathy attempt was made by the hospital staff to provide liquid food supplements and intravenous hydration.

### Lesion count methodology

An indelible marker was used by nurses to divide the patient’s body into nine different skin regions and the oropharynx when lesions involved the mouth and/or throat, for a total of 10 areas when the latter was involved. Total lesion count was determined by summing all types of lesions. Results were audited by both the Medical Monitor and study team quality control sections, utilizing the nurses case note books that included lesion count by body area.

### Clinical laboratory procedures

Clinical laboratory studies were done in compliance with USAMRIID’s Clinical Laboratory procedures. Hematology was performed using an AcT10 Hematology Analyzer by Beckman Coulter, and urinalysis was done utilizing a Multistix 10 SG, by Siemens. Clinical chemistries were performed using a Piccolo Point of Care General Chemistry panel (ALB, ALP, ALT, AMY, AST, BUN, Ca, CRE, GGT, GLU, TBIL, TP) by Abaxis. HIV Ab were done at USAMRIID using the Clinical Laboratory’s routine HIV testing kit, Bio-Rad Laboratories Multispot HIV-1/HIV-2 Rapid Test.

### DNA extraction and PCR analysis methodology

DNA extraction with QIAamp DNA Mini Kit (Qiagen), inactivation of infectious virus by incubation of specimen in extraction buffer at 56° C for 1 hr, and pan-orthopox real time polymerase chain reaction (PCR) assays were performed using a Roche LightCycler and software (24). All reagents were aliquoted into single test aliquots for up to 8 samples plus controls at USAMRIID. Each patient assay used new reagent aliquots done the day specimens were collected. Each assay included 6 standards covering the anticipated minimum and maximum values. Positive controls (EDTA blood spiked with gamma irradiated monkeypox) and negative controls (EDTA blood only) we’re extracted with each assay run for specimens and results had to fall within expected values for extraction and assay to be valid. The PCR assay included samples to determine if contamination had occurred. In cases where controls did not meet requirements tests were repeated with new reagents and standards.

### IgM and IgG methodology

Serum IgM and IgG specific against anti-OPXV were detected by ELISA as previously described (25). IgM and IgG assays were performed at 1:50 and 1:100 serum dilutions respectively using purified vaccinia virus DryVax strain. Samples were determined positive or negative based on the OD-COV value (Optical Density at 450 nm minus cutoff value of 3 times standard deviation of averages from 5 negative controls.

### Benefits to the Kole Hospital and Community

The study provided the following benefits to the Kole Hospital, staff and all patients: funding of salaries of hospital staff involved with care of MPX patients, supplementation of pharmaceutical and expendable supply budget, space available airlift of personnel and supplies, including food/nutritional support for MPX patients and their family caregivers, and funding of all medications provided to all MPX patients. The community had internet access with multiple terminals using 5% of the study internet satellite, and hospital physicians had unlimited access. Hospital clinicians had access to study clinical laboratory testing not routinely available at this hospital, as well as supplementation to the clinical staff with additional physicians to insure careful monitoring of disease and detection of treatable complications of illness.

A single payment of U.S. $20.00 was provided to study patients who returned for the follow up visit to compensate them for the blood draw. No other direct compensation was provided to study patients.

### Statistical Methods

All statistical analyses were performed using SAS version 9.4 (SAS Institute, Cary, NC) and Statistical Package R version 3.6.1. P-value less than 0.05 (typically ≤ 0.05) were statistically significant.

Study Day 0 was defined as the admission day. Patients presented at various stages of disease on the admission day. Thus for some analyses, the day of onset of rash was used to synchronize timing. Positive days were counted forward from Day 0 and negative days were counted backward from Day 0 after the rash onset. Patient age in years was computed as the integer value (nearest rounded year) based on the date of birth and the date of signed informed consent. Descriptive statistics were used to summarize the study results. PCR results and total lesion counts were log_10_ transformed for analyses. The age at admission was categorized as <5, 5-11, and ≥12 years. Repeated measurement Analysis of Variance (ANOVA) was used to compare means of total lesion count between different age groups. Repeated measurement ANOVA also was used to compare the means of blood PCR viral load between potential primary vs secondary cases or male vs female. Separate generalized estimating equations (GEE) with cumulative logit models were used to evaluate associations between: clinical symptoms/signs severity/illness severity/clinical syndromes severity (ordinal data) with total lesions, PCR results, clinical observations and vital signs; and total lesions severity (ordinal data) between vital signs, PCR results and clinical observations. Wilcoxon-Mann-Whitney test by ranks was utilized for comparison of continuous variables between two group and Kruskal Wallis Test for more than two groups. Fisher exact test or Chi-square test was used to compare categorical variables. The p value in groups were adjusted by the stepdown Bonferroni correction method.

## Results

### Patient characteristics and demographics

Two-hundred forty-four (244) patients were enrolled in the study based upon the clinical diagnosis of MPXV infection. Of these, 216 patients had MPXV infection based upon Pan-Orthopox PCR positive and MPXV specific PCR positive results. The 216 patients with active MPXV infection were monitored and provided the data for this study. The first patient was enrolled 16 March 2007 and the last patient completed the study on 02 August 2011.

Patients were predominantly male (63.9%), age range 0-61 years (mean = 14, median = 13) (Table 1). Of the 216 patients, 118 (54.6%) were age ≥ 12. Four (4) patients reported history of smallpox vaccination, born during the years 1962-1972. However, 1 of 4 did not include the presence or absence of a scar notation. One patient was confirmed HIV positive based on Ab test; once recovered from acute MPXD, he was referred to a regional hospital for HIV treatment.

**Table 1.** Demographics and exposure history

| Characteristic | Age Group |  |  | Total<br>(N 216) |
| --- | --- | --- | --- | --- |
| | < 5<br>(N = 31) | 5-11<br>(N = 67) | $\geq 12$<br>(N = 118) | |
| **Age at Admission (years), Mean (SD)** | 2 (1.3) | 8 (2.1) | 21 (8.4) | 14 (9.9) |
| **Gender, n (%)** |  |  |  |  |
| Female | 19 (61.3%) | 20 (29.9%) | 39 (33.1%) | 78 (36.1%) |
| Male | 12 (38.7%) | 47 (70.1%) | 79 (66.9%) | 138 (63.9%) |
| **Marital Status, n (%)** |  |  |  |  |
| Married | 0 (0%) | 0 (0%) | 35 (29.7%) | 35 (16.2%) |
| Single | 31 (100%) | 67 (100%) | 83 (70.3%) | 181 (83.8%) |
| **Family Exposure, n (%)** |  |  |  |  |
| Clean/Dressed Consumption Of Wild Game | 17 (54.8%) | 53 (79.1%) | 86 (72.9%) | 156 (72.2%) |
| Handled Uncooked, Freshly Butchered Meat | 14 (45.2%) | 47 (70.1%) | 72 (61.0%) | 133 (61.6%) |
| Meat Of Ground Squirrel | 8 (25.8%) | 30 (44.8%) | 50 (42.4%) | 88 (40.7%) |
| Initial Close Contact Of Infected Individual (Household) | 17 (54.8%) | 23 (34.3%) | 46 (39.0%) | 86 (39.8%) |
| Meat Of Monkey | 9 (29.0%) | 21 (31.3%) | 52 (44.1%) | 82 (38.0%) |
| Initial Mpx Contact With Blood, Body Fluids, Or Person With Tissue Or Secretions (Mpx Compatible Ill) | 12 (38.7%) | 12 (17.9%) | 33 (28.0%) | 57 (26.4%) |
| Dead Animal | 3 (9.7%) | 17 (25.4%) | 27 (22.9%) | 47 (21.8%) |
| Other Wild Game, Specify | 8 (25.8%) | 16 (23.9%) | 22 (18.6%) | 46 (21.3%) |
| Meat Of Gambian Rat Or Other Rodent | 1 (3.2%) | 5 (7.5%) | 5 (4.2%) | 11 (5.1%) |
| Multiple Exposures ($\geq 2$) | 24 (77.4%) | 67 (100%) | 114 (96.6%) | 205 (94.9%) |

### Exposure history

The most commonly reported family exposure was cleaning/dressing/consumption of wild game (n=156, 72.2%), followed by handling uncooked, freshly butchered meat (n = 133, 61.6%) and eating squirrel meat (n=88, 40.7%) (Table 1). Importantly, 205 (94.9%) patients had 2 or more possible exposures. Of the 11 individuals with a single exposure history, 4 were exposed to a household contact with MPXV infection, 4 consumed squirrel meat, 2 ate monkey meat and one cleaned, dressed, and consumed wild game (unspecified).

### Clinical symptoms

The most common clinical complaint was rash (96.8%), followed by malaise (85.2%), sore throat (78.2%), lymphadenopathy (57.4%) and anorexia (50.0%) (Table 2A). The clinical symptom occurred at least once from admission day (study day 0) to study day 3.

**Table 2A.**
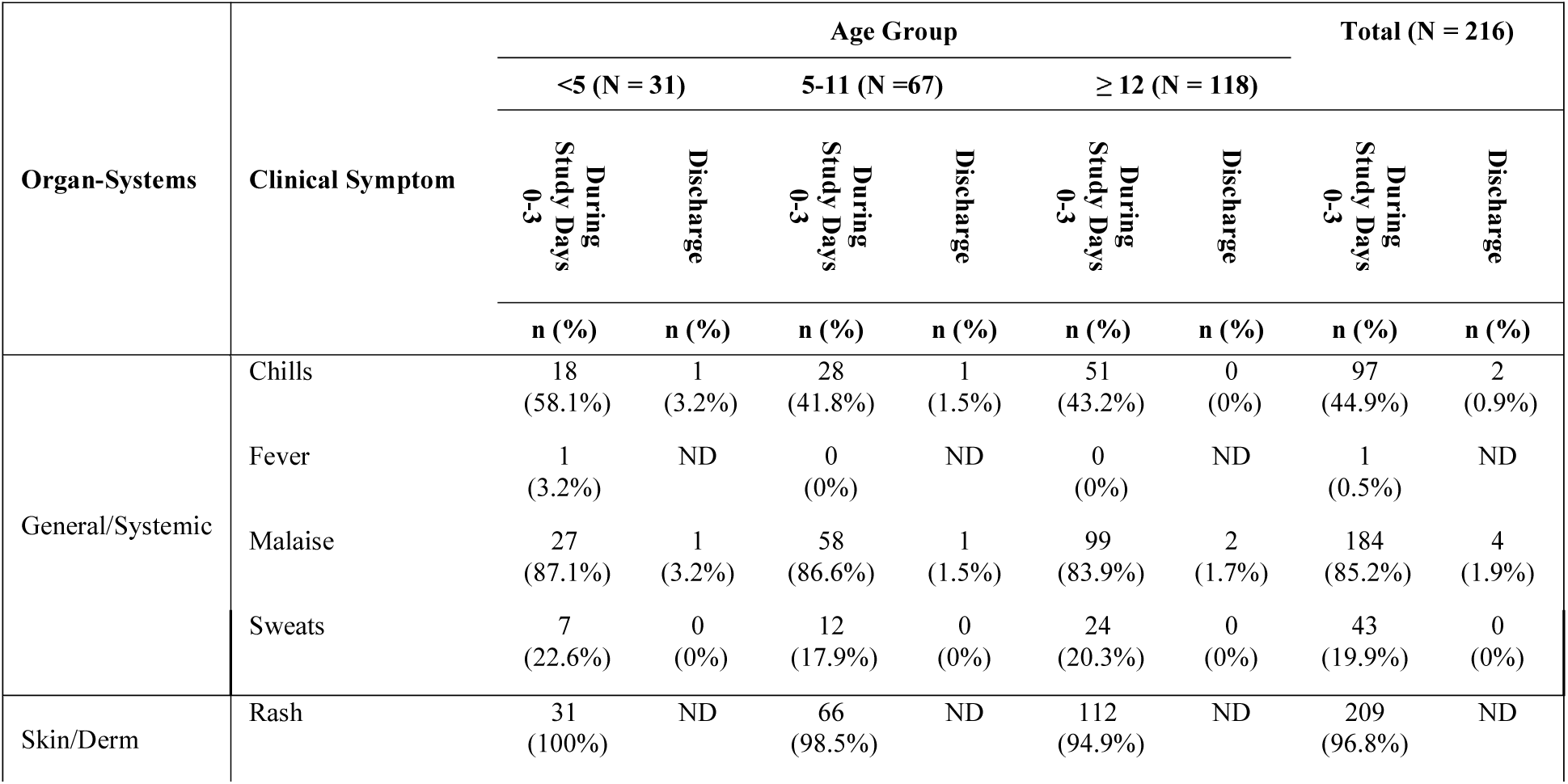
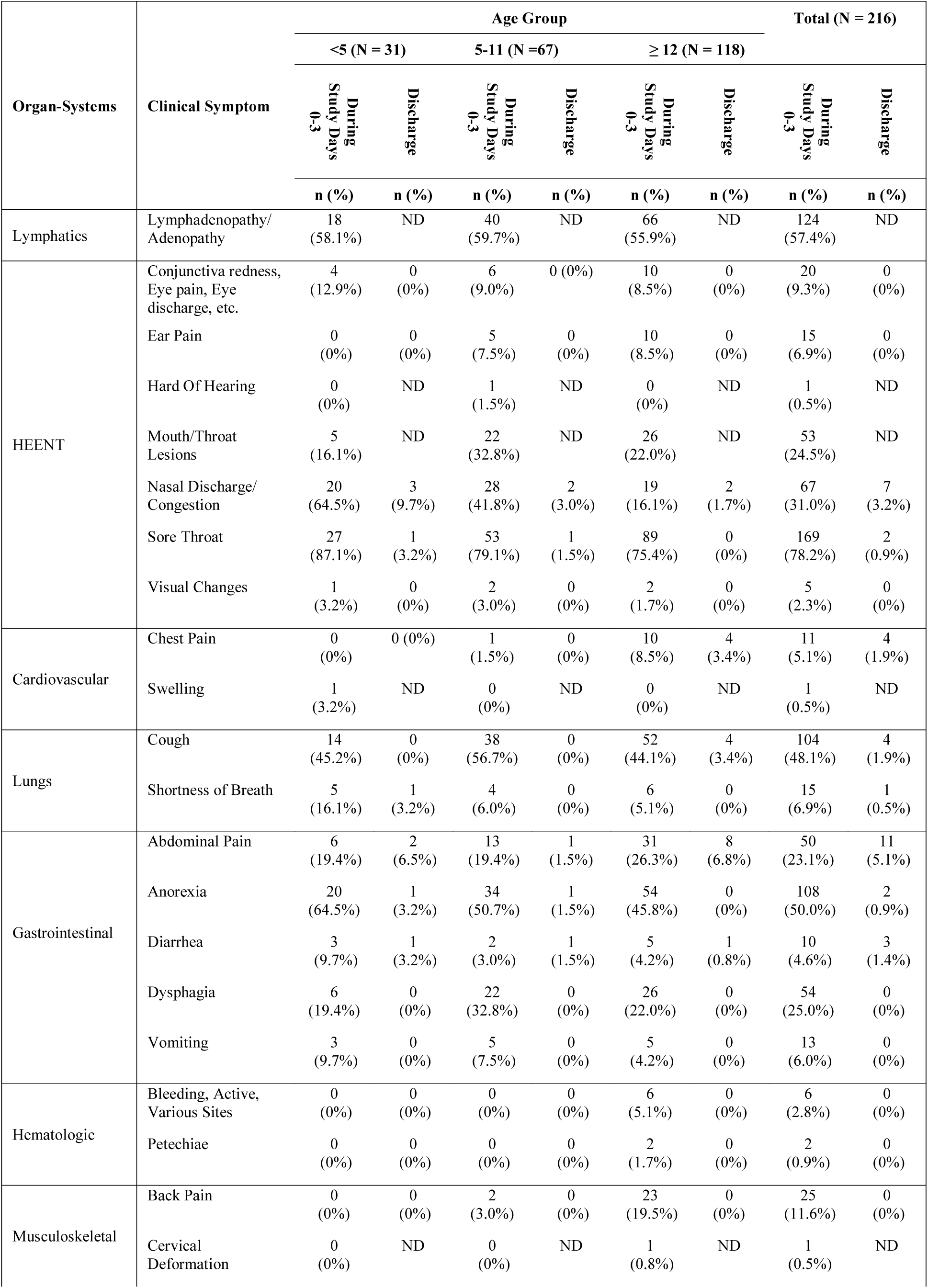
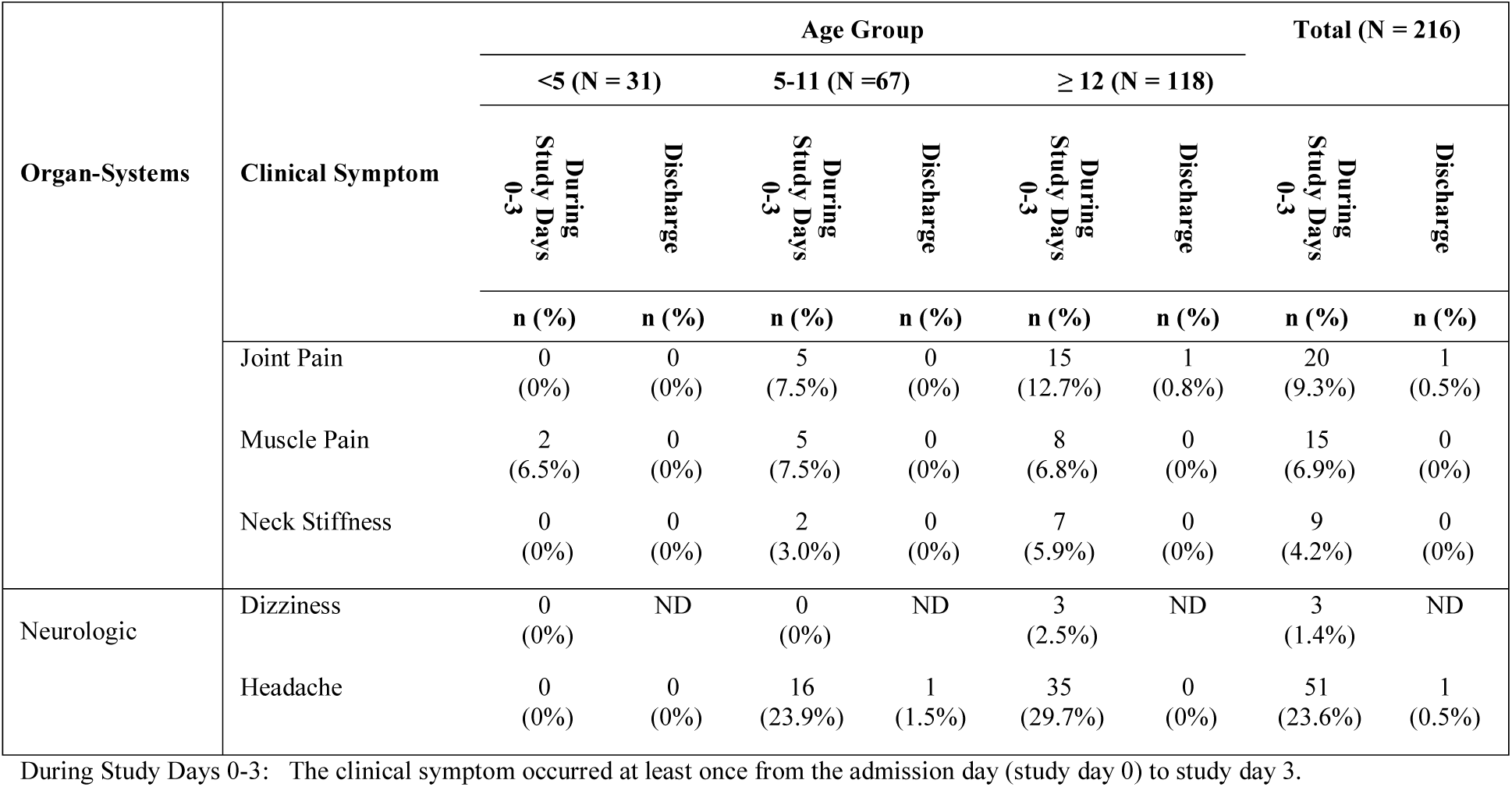
Descriptive tables of clinical symptoms in patients with acute monkeypox infection by age

**Table 2B.**
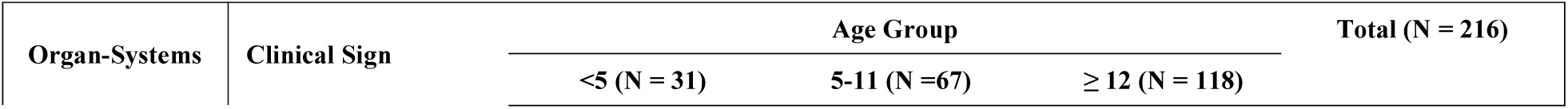
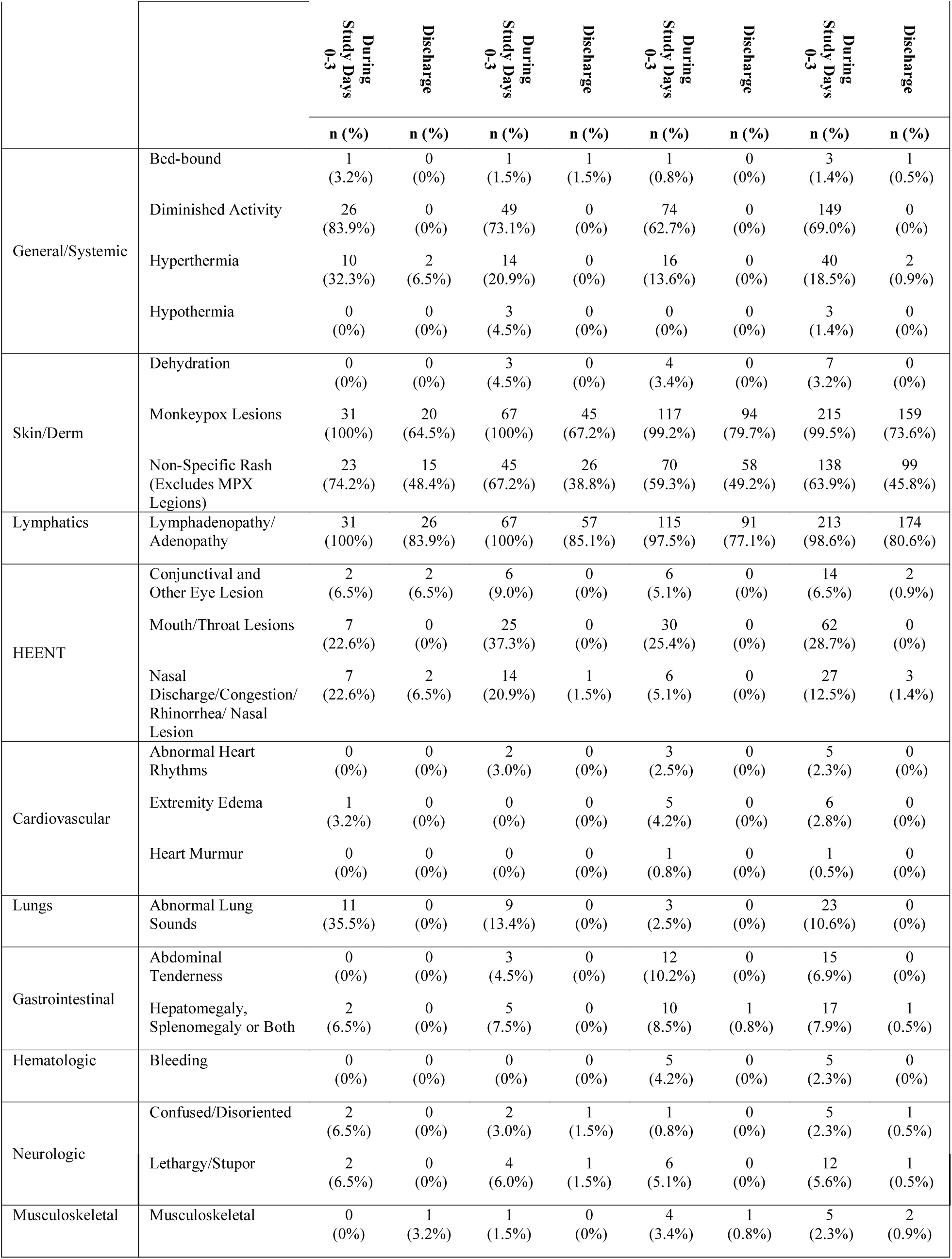
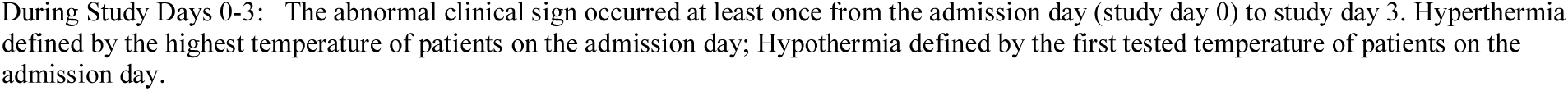
Descriptive tables of clinical signs in patients with acute monkeypox infection by age

### Physical examination findings or signs

The most common physical examination finding was classic MPXV skin lesions 215/216 (99.5%) and lymphadenopathy (adenopathy) (98.6%) (Table 2B). MPXV mouth/throat lesions were seen in 28.7%) of patients. Abnormal lung sounds were detectable in 10.6% of patients. Hepatomegaly, splenomegaly or both were noted in 7.9% of patients. Bleeding was seen in 2.3% of patients. The clinical sign occurred at least once from admission day (study day 0) to study day 3. Hyperthermia defined by the highest temperature of patients on the admission day; hypothermia defined by the first temperature of patients on the admission day.

### Duration of symptoms and signs

Figure 1 shows the duration of symptoms and signs (S&S) from admission day in patients with acute disease. As expected, at day of discharge the majority-of-patients still had evidence of monkeypox skin lesions and lymphadenopathy. Most S&S lasted 3-5 days. The sequence of symptom onset was not easily determined by this dataset.

**Figure 1.**
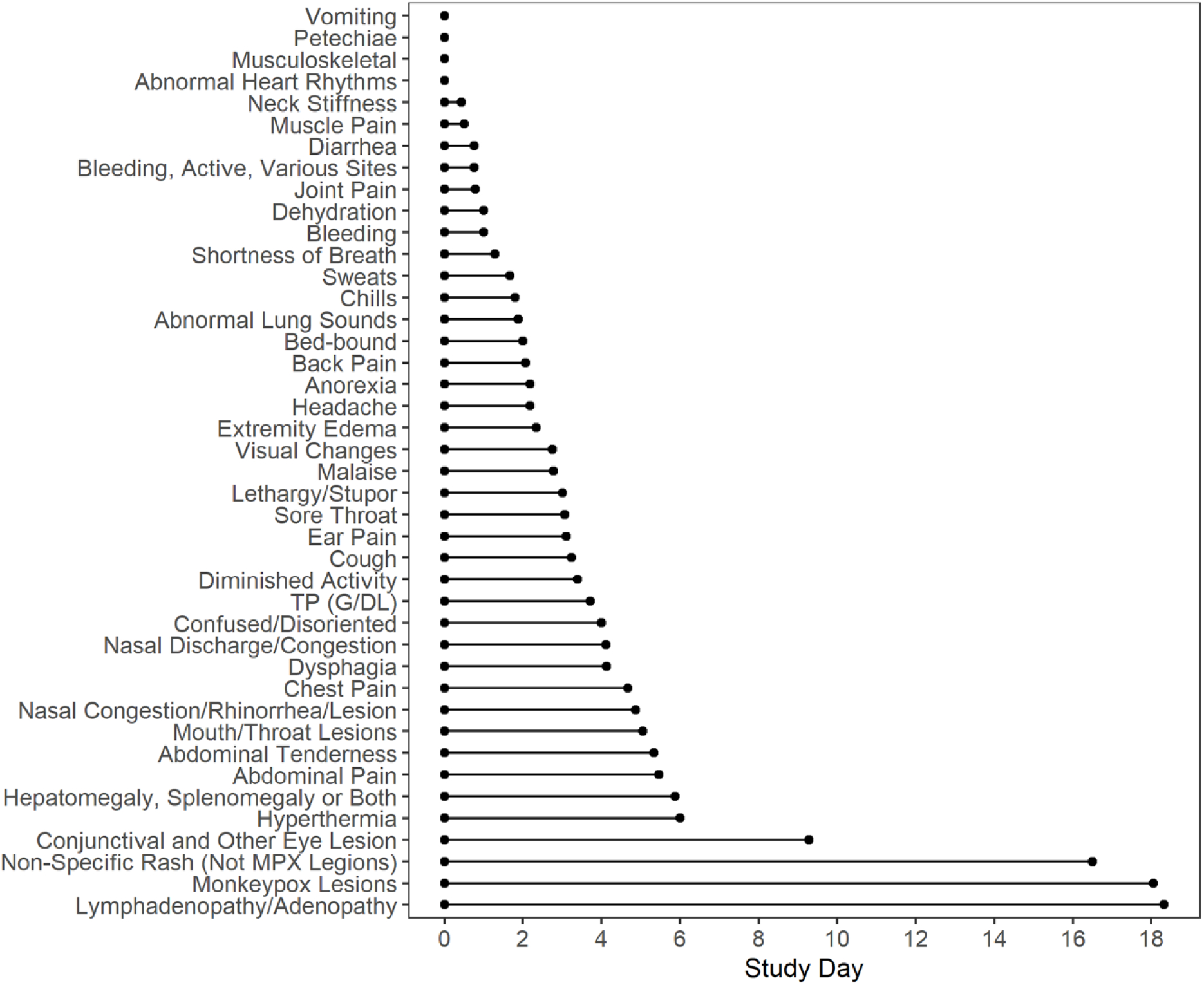
The duration of clinical symptoms and signs. The mean of duration only included clinical symptoms and clinical signs present on the admission day.

### Disease severity categories

Clinical illness groups are based upon the numbers of clinical S&S occurring on admission as outlined at Supplemental Table S1. The matrix at Figure 2 shows the fitness of clinical illness categories based upon clinical S&S severity.

**Figure 2.**
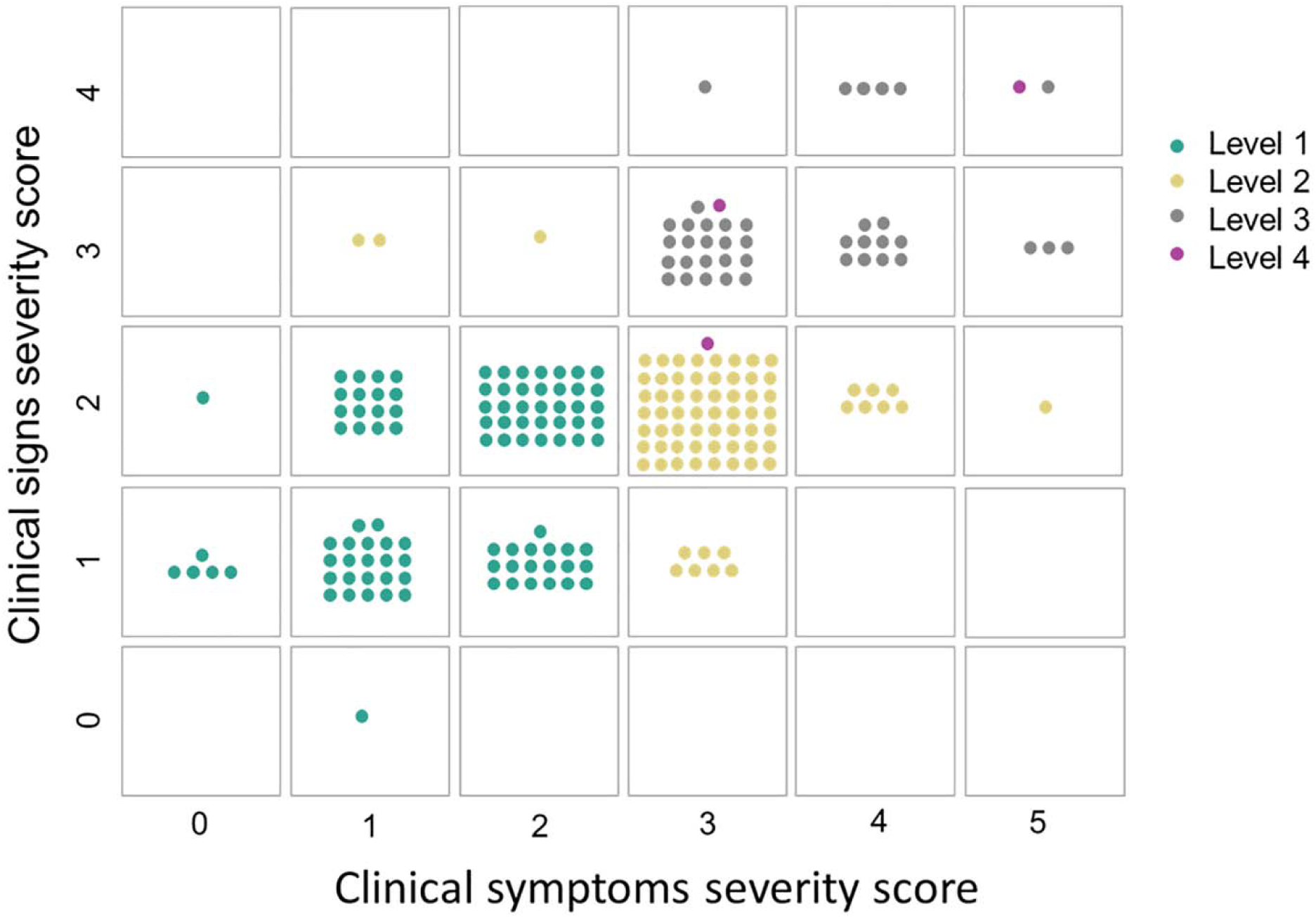
Defining Monkeypox illness severity: clinical symptoms severity score and clinical signs severity score matrix. Clinical symptom/sign severity was defined as the number of any clinical symptom/sign during study day 0-3. Level 4 is the fatal group and the patient (cell 3-2) in this group was less than 1 year old with limited ability to explain his clinical symptoms.

The disease illness categories are: *level 1, level 2, level 3, and level 4 (fatal)*. We were not able to delineate S&S that define survival versus death due to low statistical power. However, we did find a group of S&S that defined the level 1, level 2, level 3 and level 4 (fatal) (Table 3A and 3B) based upon the frequency of the S&S. Common symptoms included sore throat, anorexia, cough, chills, nasal discharge & congestion, dysphagia, mouth/throat lesions, headache, abdominal pain, sweats conjunctiva lesions, shortness of breath as well as such physical findings or signs as diminished activity, nonspecific rash, mouth/throat lesions, abnormal lung sounds, hepatomegaly/splenomegaly, lethargy/stupor, dehydration, and confusion/disorientation.

**Table 3A.** Comparison of clinical symptoms among monkeypox illness severity categories

| Organ-Systems | Clinical Symptom | Level 1<br>(N = 99) | Level 2<br>(N = 74) | Level 3<br>(N = 40) | Adjusted<br>P value | Raw P<br>value |
| --- | --- | --- | --- | --- | --- | --- |
|  |  | n (%) | n (%) | n (%) |  |  |
| General | Chills | 18 (18.2) | 46 (62.2) | 31 (77.5) | <.0001 | <.0001 |
|  | Malaise | 74 (74.7) | 68 (91.9) | 39 (97.5) | 0.0043 | 0.0003 |
|  | Sweats | 7 (7.1) | 18 (24.3) | 17 (42.5) | <.0001 | <.0001 |
| Skin/Derm | Rash | 94 (94.9) | 73 (98.6) | 39 (97.5) | 0.8391 | 0.4195 |
| Lymphatics | Lymphadenopathy/Adenopathy | 54 (54.5) | 40 (54.1) | 28 (70.0) | 0.6044 | 0.2015 |
| HEENT | Conjunctiva redness, Eye pain, Eye discharge, etc. | 3 (3.0) | 6 (8.1) | 11 (27.5) | 0.0019 | 0.0001 |
|  | Ear Pain | 1 (1.0) | 6 (8.1) | 7 (17.5) | 0.0095 | 0.0008 |
|  | Mouth/Throat Lesions | 8 (8.1) | 20 (27.0) | 24 (60.0) | <.0001 | <.0001 |
|  | Nasal Discharge/Congestion | 13 (13.1) | 25 (33.8) | 27 (67.5) | <.0001 | <.0001 |
|  | Sore Throat | 60 (60.6) | 68 (91.9) | 38 (95.0) | <.0001 | <.0001 |
|  | Visual Changes | 2 (2.0) | 0 (0) | 3 (7.5) | 0.2198 | 0.0440 |
| Cardiovascular | Chest Pain | 0 (0) | 7 (9.5) | 4 (10.0) | 0.0139 | 0.0014 |
| Lungs | Cough | 28 (28.3) | 47 (63.5) | 27 (67.5) | <.0001 | <.0001 |
|  | Shortness of Breath | 0 (0) | 6 (8.1) | 7 (17.5) | 0.0015 | <.0001 |
| Gastrointestinal | Abdominal Pain | 7 (7.1) | 32 (43.2) | 9 (22.5) | <.0001 | <.0001 |
|  | Anorexia | 22 (22.2) | 51 (68.9) | 32 (80.0) | <.0001 | <.0001 |
|  | Diarrhea | 2 (2.0) | 7 (9.5) | 1 (2.5) | 0.2663 | 0.0666 |
|  | Dysphagia | 3 (3.0) | 21 (28.4) | 29 (72.5) | <.0001 | <.0001 |
|  | Vomiting | 1 (1.0) | 7 (9.5) | 4 (10.0) | 0.0919 | 0.0115 |
| Hematologic | Bleeding, Active, Various Sites | 1 (1.0) | 1 (1.4) | 4 (10.0) | 0.1708 | 0.0244 |
|  | Petechiae | 0 (0) | 0 (0) | 2 (5.0) | 0.2073 | 0.0345 |
| Musculoskeletal | Back Pain | 5 (5.1) | 17 (23.0) | 3 (7.5) | 0.0133 | 0.0012 |
|  | Joint Pain | 3 (3.0) | 11 (14.9) | 6 (15.0) | 0.0607 | 0.0067 |
|  | Muscle Pain | 0 (0) | 10 (13.5) | 5 (12.5) | 0.0019 | 0.0001 |
|  | Neck Stiffness | 0 (0) | 3 (4.1) | 6 (15.0) | 0.0043 | 0.0003 |
| Neurologic | Dizziness | 1 (1.0) | 1 (1.4) | 1 (2.5) | 0.8391 | 0.7739 |
|  | Headache | 7 (7.1) | 31 (41.9) | 11 (27.5) | <.0001 | <.0001 |

Statistics performed by Fisher exact test or Chi-square test with stepdown Bonferroni correction. Significant differences are highlighted by yellow (adjusted $p$ value $\leq 0.05$ )

**Table 3B.** Comparison of clinical signs among monkeypox illness severity categories

| Organ-Systems | Clinical Sign | Level 1<br>(N = 99) | Level 2<br>(N = 74) | Level 3<br>(N = 40) | Adjusted<br>P value | Raw P<br>value |
| --- | --- | --- | --- | --- | --- | --- |
|  |  | n (%) | n (%) | n (%) |  |  |
| General/Systemic | Bed-bound | 1 (1.0) | 0 (0) | 2 (5.0) | 0.3991 | 0.1320 |
|  | Diminished Activity | 47 (47.5) | 61 (82.4) | 38 (95.0) | <.0001 | <.0001 |
|  | Hyperthermia | 5 (5.1) | 20 (27.0) | 13 (32.5) | 0.0001 | <.0001 |
|  | Hypothermia | 1 (1.0) | 1 (1.4) | 1 (2.5) | 1.0000 | 0.7739 |
| Skin/Derm. | Dehydration | 0 (0) | 1 (1.4) | 6 (15.0) | 0.0010 | <.0001 |
|  | Non-Specific Rash (Excludes MPX Lesions) | 72 (72.7) | 46 (62.2) | 17 (42.5) | 0.0328 | 0.0041 |
| Lymphatics | Lymphadenopathy/Adenopathy | 98 (99.0) | 73 (98.6) | 39 (97.5) | 1.0000 | 0.7739 |
| HEENT | Conjunctival and Other Eye Lesion | 3 (3.0) | 4 (5.4) | 7 (17.5) | 0.0659 | 0.0110 |
|  | Mouth/Throat Lesions | 9 (9.1) | 23 (31.1) | 29 (72.5) | <.0001 | <.0001 |
|  | Nasal Discharge/Congestion/Rhinorrhea/Nasal Lesion | 2 (2.0) | 5 (6.8) | 19 (47.5) | <.0001 | <.0001 |
| Cardiovascular | Abnormal Heart Rhythms | 1 (1.0) | 0 (0) | 3 (7.5) | 0.1080 | 0.0216 |
|  | Extremity Edema | 1 (1.0) | 0 (0) | 5 (12.5) | 0.0107 | 0.0010 |
| Lungs | Abnormal Lung Sounds | 2 (2.0) | 10 (13.5) | 11 (27.5) | 0.0003 | <.0001 |
| Gastrointestinal | Abdominal Tenderness | 1 (1.0) | 8 (10.8) | 6 (15.0) | 0.0118 | 0.0013 |
|  | Hepatomegaly, Splenomegaly or Both | 2 (2.0) | 4 (5.4) | 11 (27.5) | 0.0003 | <.0001 |
| Hematologic | Bleeding | 1 (1.0) | 1 (1.4) | 3 (7.5) | 0.3991 | 0.0998 |
| Neurologic | Confused/Disoriented | 0 (0) | 0 (0) | 4 (10.0) | 0.0110 | 0.0011 |
|  | Lethargy/Stupor | 1 (1.0) | 1 (1.4) | 9 (22.5) | <.0001 | <.0001 |
| Musculoskeletal | Musculoskeletal | 1 (1.0) | 0 (0) | 4 (10.0) | 0.0331 | 0.0047 |

Statistics performed by Fisher exact test or Chi-square test with stepdown Bonferroni correction. Significant differences are highlighted by yellow (adjusted $p$ value $\leq 0.05$ )

### Monkeypox rash characteristics and body region distribution

Figure 3 depicts the monkeypox lesion distribution pattern graphically. The graphic shows the mean distribution of all patients at peak lesion count (photographs of oropharyngeal lesions are cropped-consult authors for full de-identifiable photographs). Figure 4A shows the mean (±SE) total lesion count by age group and time from day of onset of classic MPXV rash. The mean lesion count was higher for the group <5 years compared to age groups 5-11 and ≥12 years, although the difference was not statistically significant (p = 0.1630). Lesions peaked between days 5-8.

**Figure 3.**
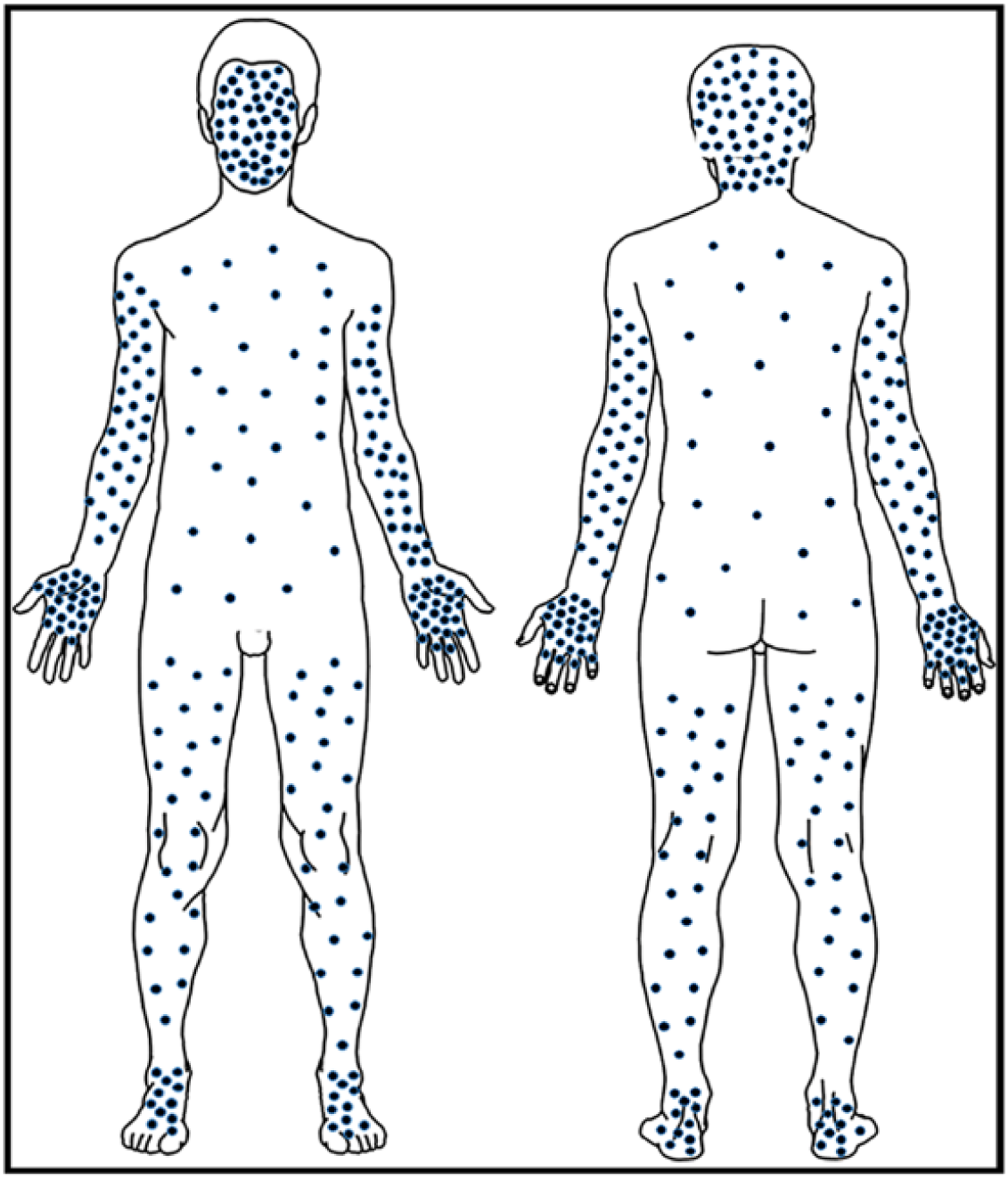
Pattern of distribution of monkeypox lesions. The pattern of distribution of monkeypox lesions. The mean of lesions were counted on the day which the patients had the highest lesion count. One dot represents about 10 lesions per area (mean of lesion count/ percentage of Body Surface Area).

**Figure 4.**
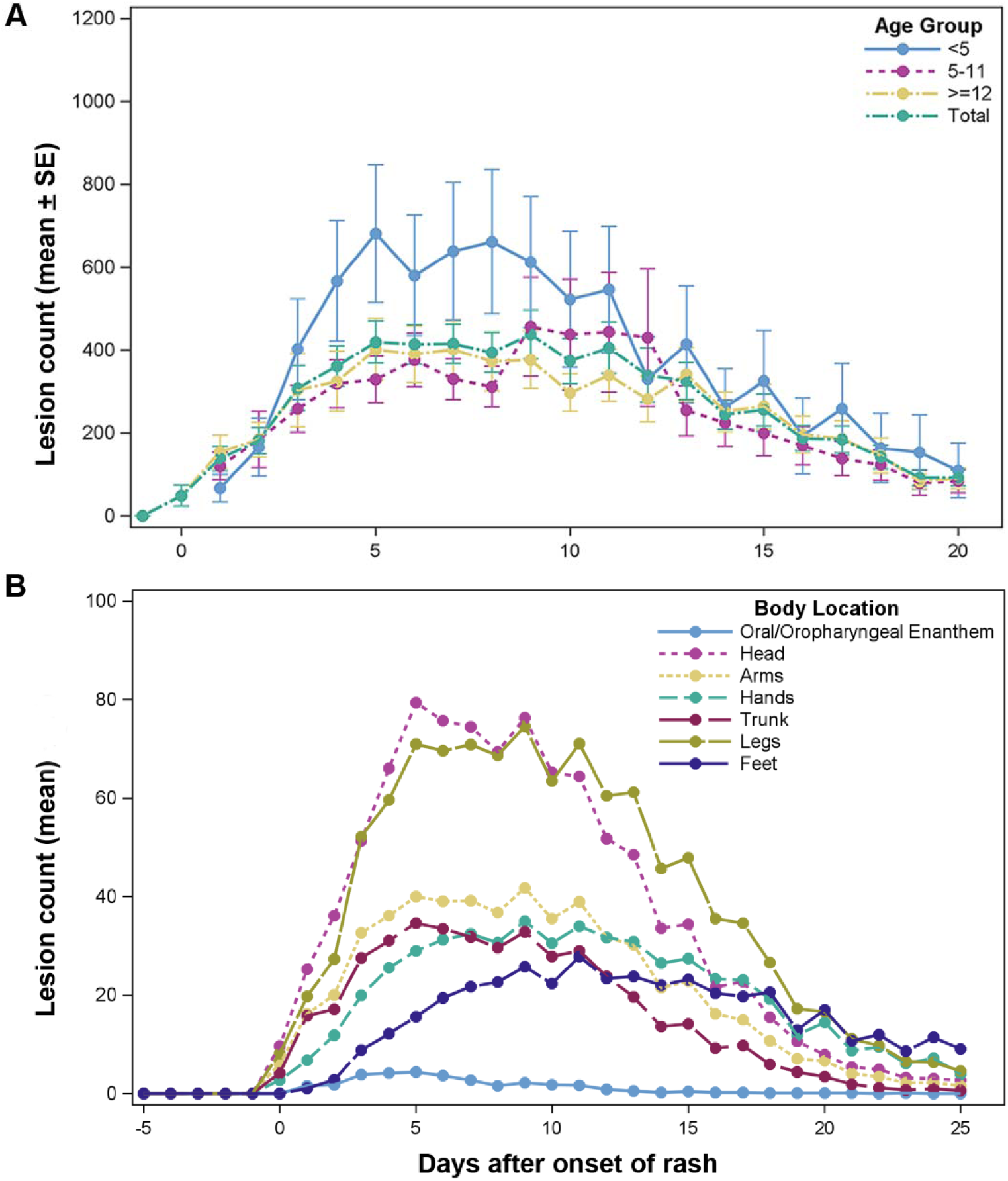
Change in total lesion count or lesion count by body location over time. (A) The mean with standard error bars of total lesion count by age group. Repeated measurement Analysis of Variance (ANOVA) showed there was no significant difference among the groups (*p* = 0.2552). (B) Distribution of mean of lesion counts for all patients by body location over time.

Lesion counts by body region over time is shown in Figure 4B. Lesion count peaked first on the head on Day 5. Figure 5 shows lesion progression from macule, papule, vesicle, pustule, umbilication, scabbing and desquamation for the hand. Progression from one phase to another occurs in order previously documented. Classic dogma holds that lesions in the same body region progress together as illustrated here.

**Figure 5.**
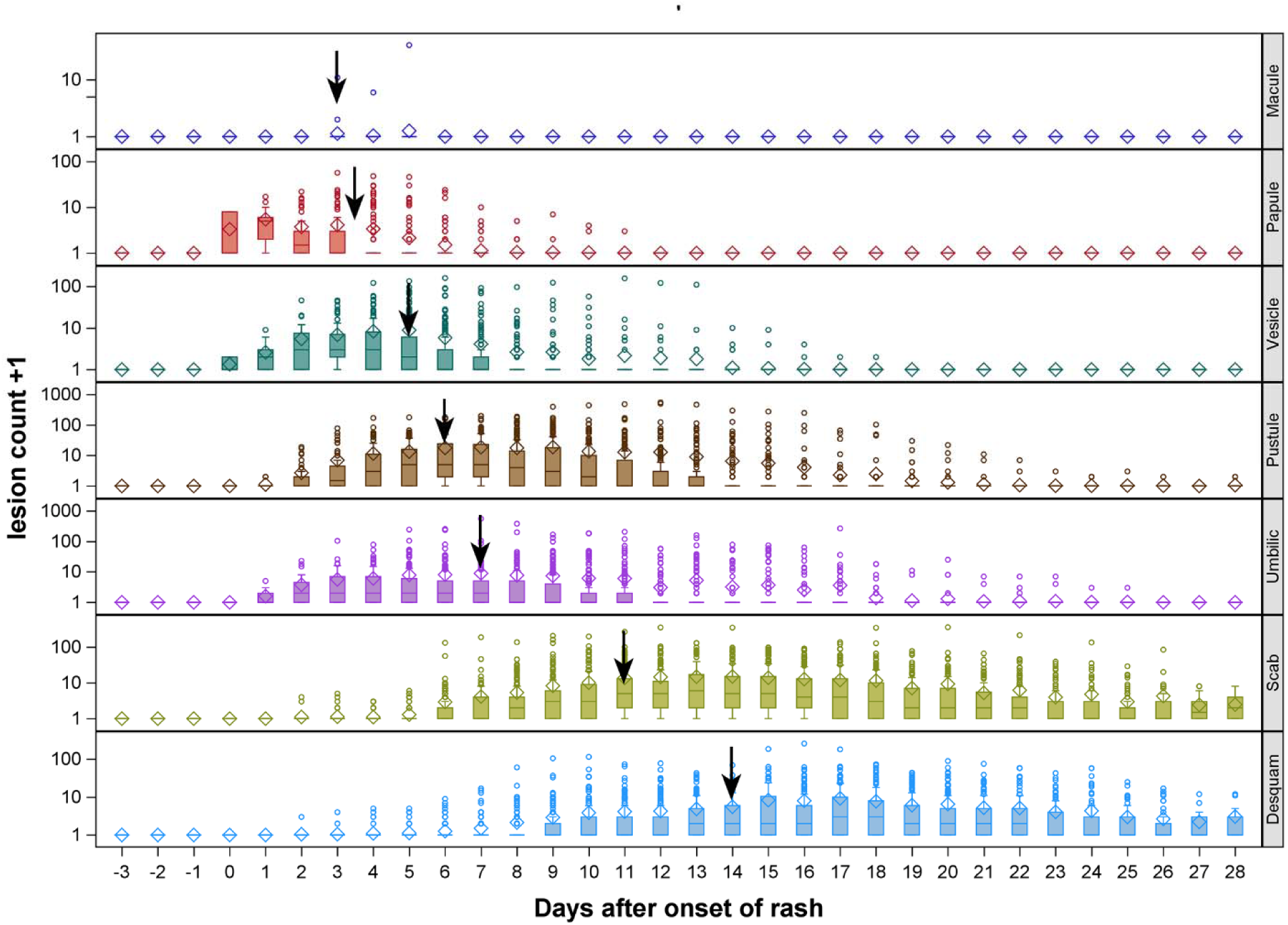
Change in lesion count on hands with time. Lesion progression from macule, papule, vesicle, pustule, umbilication (Umbilic), scabbing (scab) and desquamation (desquam). ◊ represent mean of count, ─ represent median of count, □ represent outliers, ↓ represent median day after onset of rash to get max lesion count for that specific rash morphology on hand.

Whether on day of admission or at maximum lesion count, a strong statistical significance was noted between illness categories and lesion counts independent of body location of lesions (Table 4A & B).

**Table 4A.** Comparison of lesion count by location on day of admission among illness severity categories

| Body Location | Level 1<br>(N = 99) | Level 2<br>(N = 74) | Level 3<br>(N = 40) | Level 4<br>(Death)<br>(N = 3) | Adjusted<br>P value | Raw P<br>value |
| --- | --- | --- | --- | --- | --- | --- |
|  | Mean (SD) | Mean (SD) | Mean (SD) | Mean (SD) |  |  |
| Oral/Oropharyngeal Enanthem | 1 (2.0) | 4 (10.0) | 6 (10.6) | 29 (35.9) | <.0001 | <.0001 |
| Head | 32 (44.3) | 87 (113.1) | 96 (117.6) | 191 (102.2) | <.0001 | <.0001 |
| Arms | 35 (54.0) | 89 (100.6) | 118 (181.0) | 259 (89.8) | <.0001 | <.0001 |
| Hands | 15 (32.7) | 32 (43.5) | 49 (101.7) | 73 (58.2) | <.0001 | <.0001 |
| Trunk | 27 (43.0) | 77 (89.8) | 95 (138.0) | 313 (157.1) | <.0001 | <.0001 |
| Legs | 62 (111.1) | 154 (204.4) | 217 (372.7) | 374 (343.5) | <.0001 | <.0001 |
| Feet | 12 (42.4) | 17 (28.8) | 22 (43.9) | 49 (70.7) | 0.0110 | 0.0110 |
| Total Body | 184 (300.9) | 459 (547.1) | 602 (885.8) | 1288 (763.7) | <.0001 | <.0001 |

Statistics performed by Kruskal Wallis Test with stepdown Bonferroni correction. Significant differences were highlighted by yellow (adjusted *p* value ≤ 0.05)

**Table 4B.**
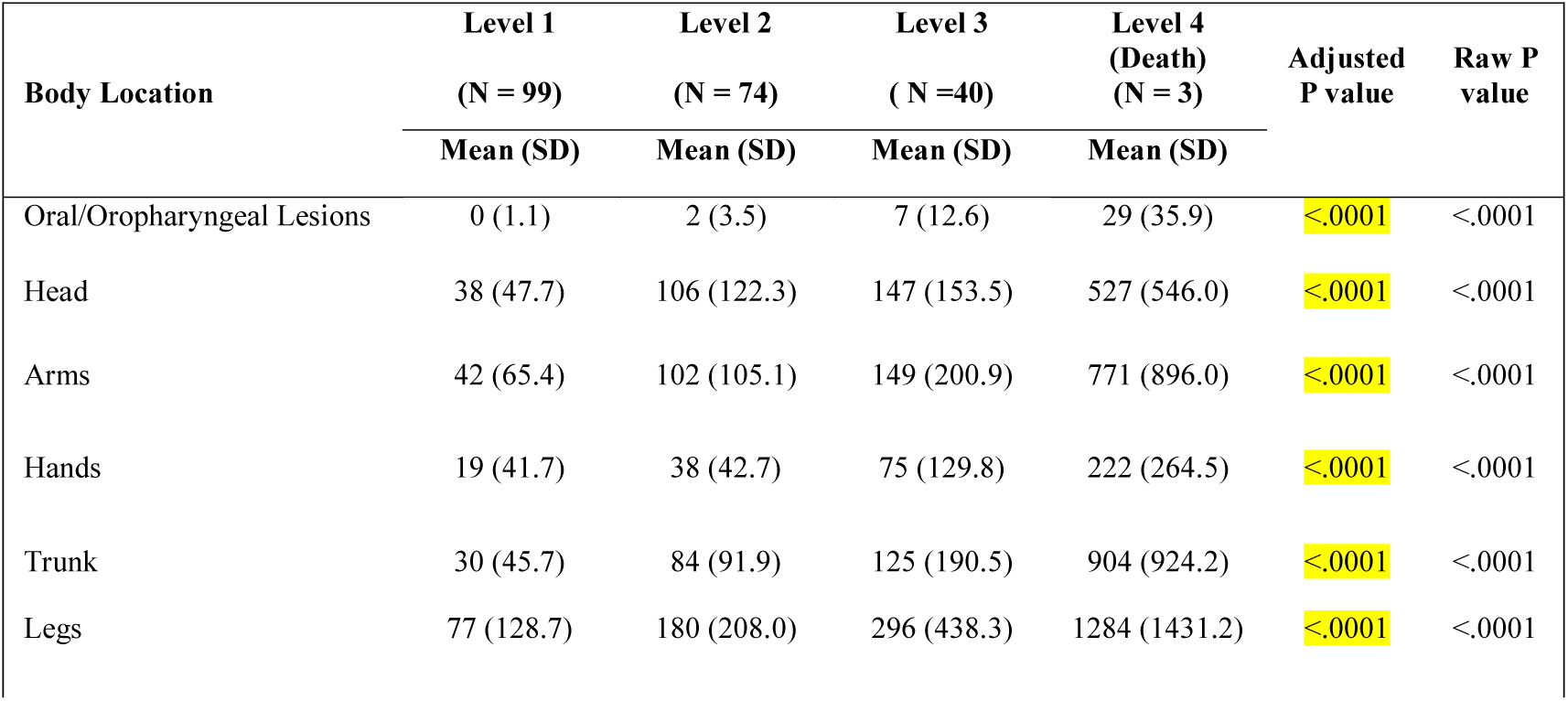
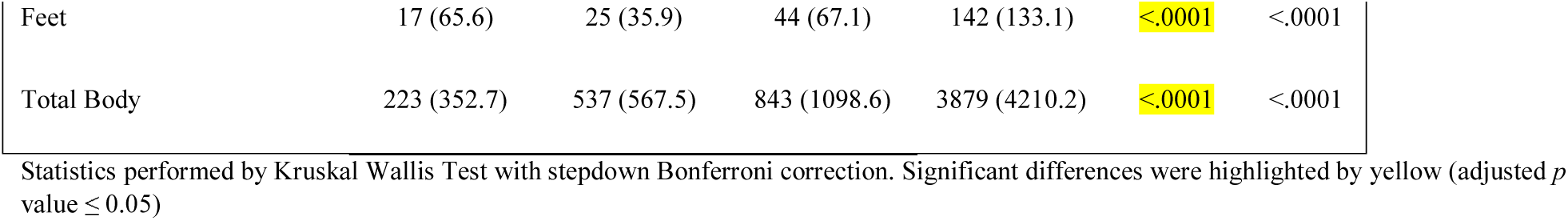
Comparison of maximum lesion count by location among illness severity categories

| Body Location | Level 1<br>(N = 99) | Level 2<br>(N = 74) | Level 3<br>(N = 40) | Level 4<br>(Death)<br>(N = 3) | Adjusted<br>P value | Raw P<br>value |
| --- | --- | --- | --- | --- | --- | --- |
|  | Mean (SD) | Mean (SD) | Mean (SD) | Mean (SD) |  |  |
| Oral/Oropharyngeal Lesions | 0 (1.1) | 2 (3.5) | 7 (12.6) | 29 (35.9) | <.0001 | <.0001 |
| Head | 38 (47.7) | 106 (122.3) | 147 (153.5) | 527 (546.0) | <.0001 | <.0001 |
| Arms | 42 (65.4) | 102 (105.1) | 149 (200.9) | 771 (896.0) | <.0001 | <.0001 |
| Hands | 19 (41.7) | 38 (42.7) | 75 (129.8) | 222 (264.5) | <.0001 | <.0001 |
| Trunk | 30 (45.7) | 84 (91.9) | 125 (190.5) | 904 (924.2) | <.0001 | <.0001 |
| Legs | 77 (128.7) | 180 (208.0) | 296 (438.3) | 1284 (1431.2) | <.0001 | <.0001 |
| Feet | 17 (65.6) | 25 (35.9) | 44 (67.1) | 142 (133.1) | <.0001 | <.0001 |
| Total Body | 223 (352.7) | 537 (567.5) | 843 (1098.6) | 3879 (4210.2) | <.0001 | <.0001 |

Statistics performed by Kruskal Wallis Test with stepdown Bonferroni correction. Significant differences were highlighted by yellow (adjusted $p$ value $\leq 0.05$ )

### Monkeypox infection associated lymphadenopathy

The frequency of monkeypox induced lymphoadenopathy was 98.6%, second only to the frequency of the classic monkeypox rash itself. The distribution of lymphoadenopathy is depicted in Figure 6 (photographs are cropped-consult authors for full de-identifiable photographs). The cervical region was most frequently afflicted at 85.6%; the second most frequent area was the inguinal region 77.3%.

**Figure 6.**
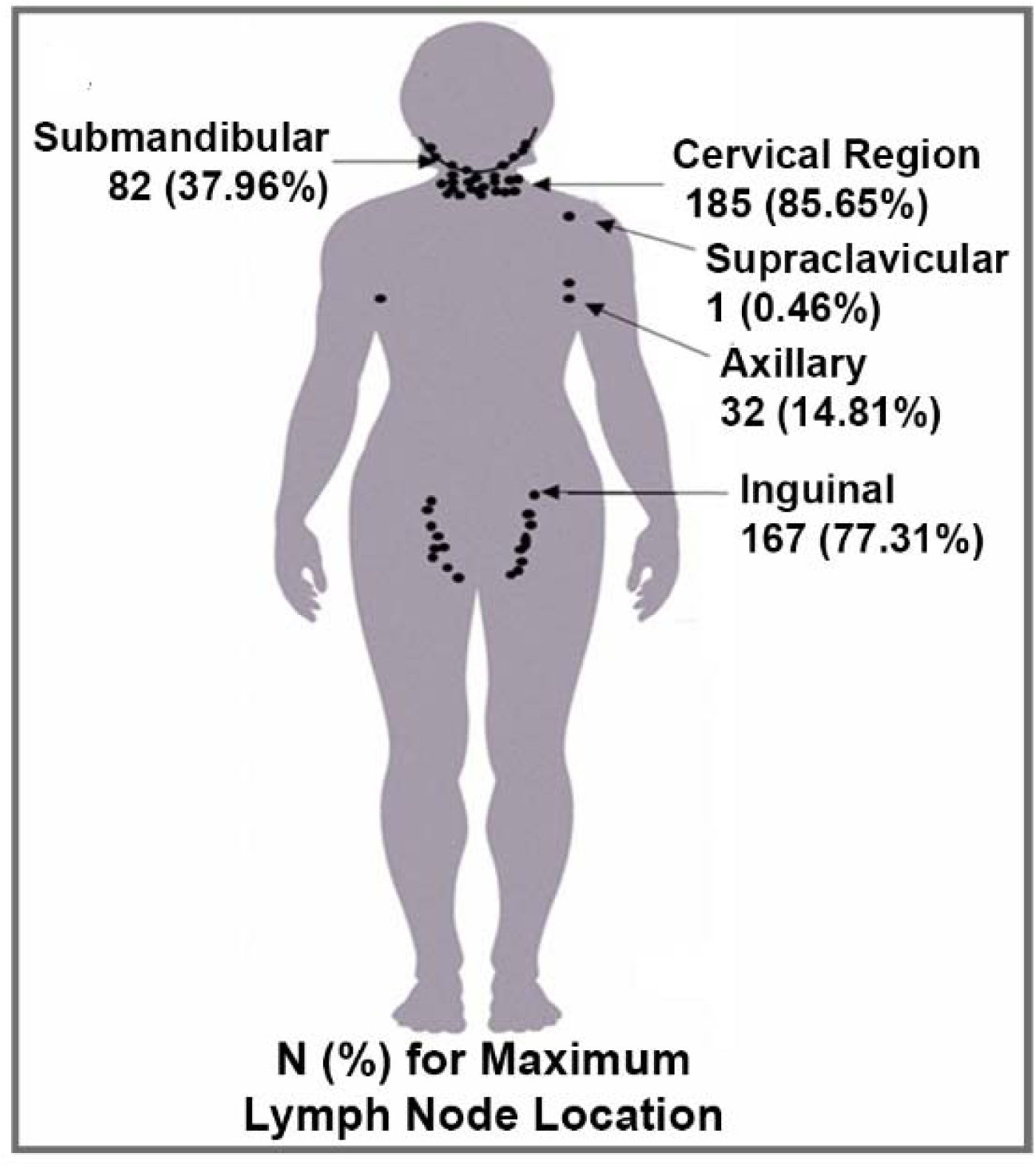
The pattern of distribution of Monkeypox associated lymph node. Monkeypox associated lymph node distribution. The lymph node count and distribution were documented on the day patients had the most lymph nodes. One dot equals 10 lymph nodes or fraction thereof depending upon the count.

### Clinical laboratory findings

The mean, median and range of values for each clinical laboratory tests are shown in Supplemental Table S2. Figure 7A & 7B compares the median values of the 3 illness severity categories-survivors--(death not included) and compares survivors (levels 1-3) vs. level 4 (death). Comparing those who died and survivors show statistically significant differences in the alanine phosphatase (ALT) (90 vs 26 U/L; p = 0.0224, adjusted) and aspartate aminotransferase (AST) (415 vs 48 U/L; p = 0.0004, adjusted). For CBC (complete blood count) variables, there were no difference between survivors and fatal cases for any CBC variable. However, the WBC and Neutrophil count show difference among the non-fatal categories. The platelet count was 130 x 10^3^/µL in the fatal group vs 296 x 10^3^/µL among survivors (p = 0.0102, unadjusted). For urine, the only statistically significant finding was elevated protein among illness severity level 1, 59 mg/dL (SD 65.3), level 2 category 88 mg/dL (SD 87.3), vs level 3 category 114 mg/dL (SD 108.3); p = 0.0147, adjusted (data not shown).

**Figure 7A.**
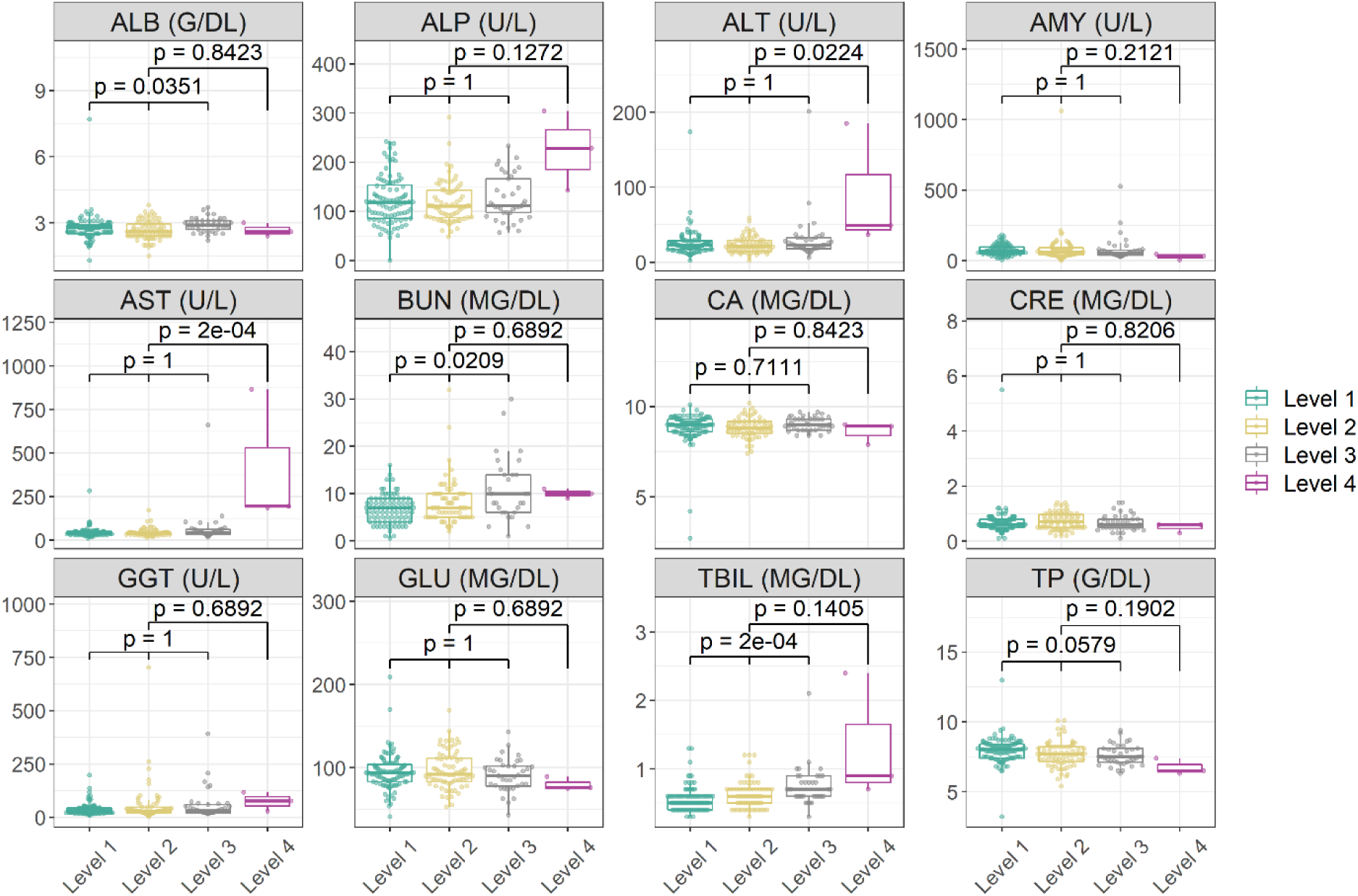
Comparison of clinical laboratory among monkeypox illness severity categories. Comparison of clinical laboratory tests of patients with active monkeypox illness by severity categories on admission day. Statistics performed by Wilcoxon-Mann-Whitney test by ranks or Kruskal Wallis Test with stepdown Bonferroni correction. □ represent the first quartile to the third quartile of value in the group, ─ represent median value in the group, ○ represent each value in the group.

**Figure 7B.**
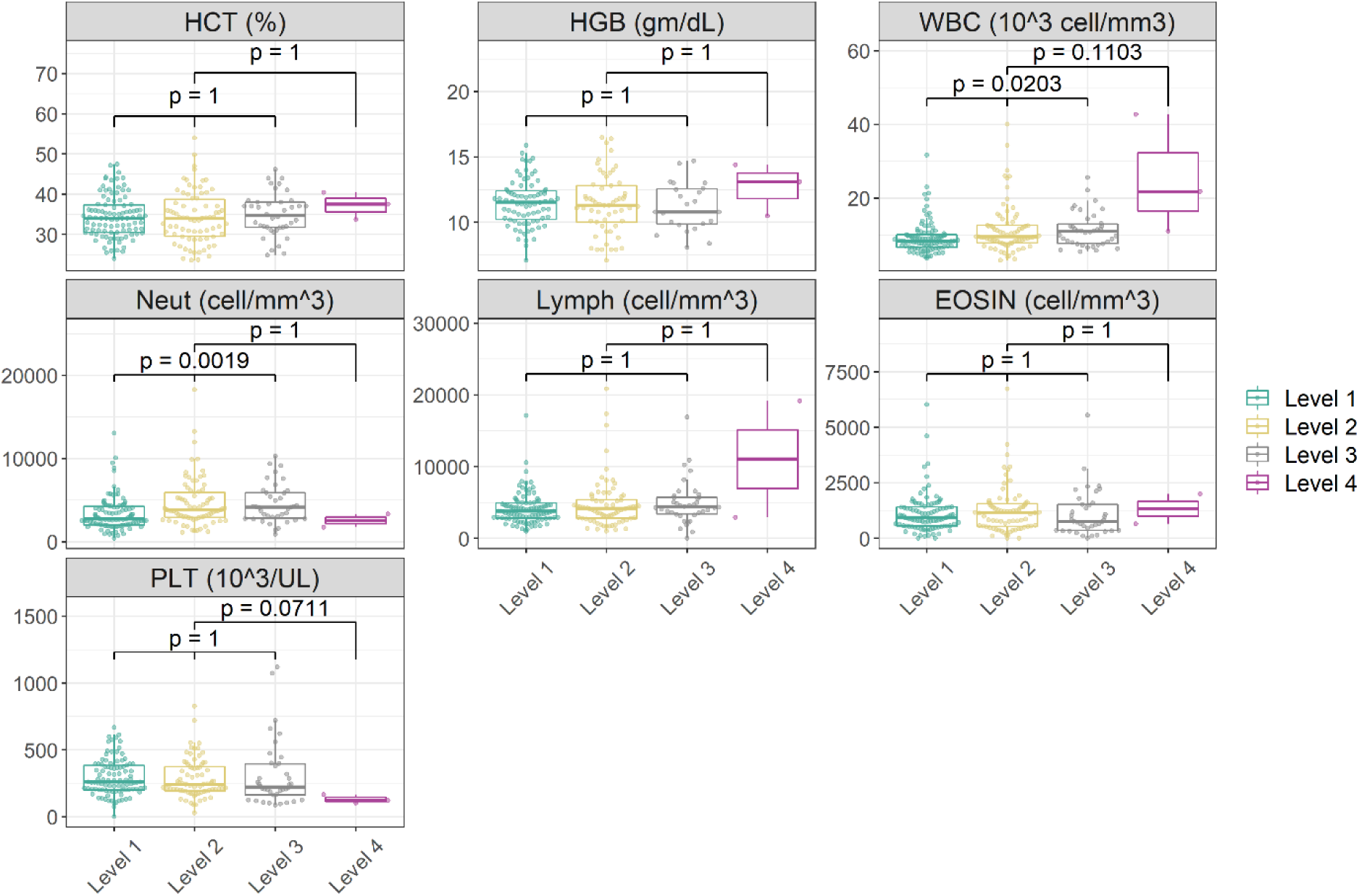
Comparison of CBC in patients with monkeypox illness severity categories. Comparison of complete blood count (CBC) of patients with active monkeypox illness by severity categories on the admission day. Statistics performed by Wilcoxon-Mann-Whitney test by ranks or Kruskal Wallis Test with stepdown Bonferroni correction. □ represent the first quartile to the third quartile of value in the group, ─ represent median value in the group, ○ represent each value in the group.

### IgM and IgG antibody responses

A total of 200 patients serum samples were tested for IgM and IgG by ELISA. A total of 189 (94.5%) develop IgM responses and all 200 (100%) were either IgG positive at enrollment or became positive during their hospitalization. The results of GEE with a cumulative logit model showed that total lesion severity was not significantly associated with IgG antibody responses (OR = 1.38, 95% CI: 0.72-2.62, p = 0.3597). However, the same model showed IgM antibody responders were 5.09 times more likely to have higher lesion severity association than IgM non-responders (OR = 5.09, 95% CI: 2.91-8.93, p < 0.0001). Further analyses of these individuals will be the topic of a separate report.

### PCR results

No significant change in MPX virus occurred between 1979 and 2010 based on full length sequence of MPX from a patient. There were only 17 base changes out of 186,000 base pairs between Zaire 79 and 136 even though they were isolated 30 years apart.

Blood, throat swabs, skin lesion and scab specimens were collected at scheduled intervals per protocol to the follow up clinical visit at Day 75 and tested by PCR for confirmation of MPXV infections. Figure 8 shows MPXV genomic material detection in blood and pharyngeal swabs by PCR occurs before the onset of the classic rash onset. Therefore, care should be taken when comparing PCR results with other variables because the maximum PCR viral load in blood occurs near the first day of rash appearance, often before patients present to the hospital. Unlike maximum lesion count maximum blood PCR viral load often could not be accurately determined in this study—by the time most patients arrived at the hospital, viral load would have already peaked. The graphs show a decrease in viral load over time for blood (p < 0.0001) and pharynx (p < 0.0001). Generally, for specimens collected at the same time from the same patient, the PCR viral load from throat is about 2000 genomes/mL higher than blood.

**Figure 8.**
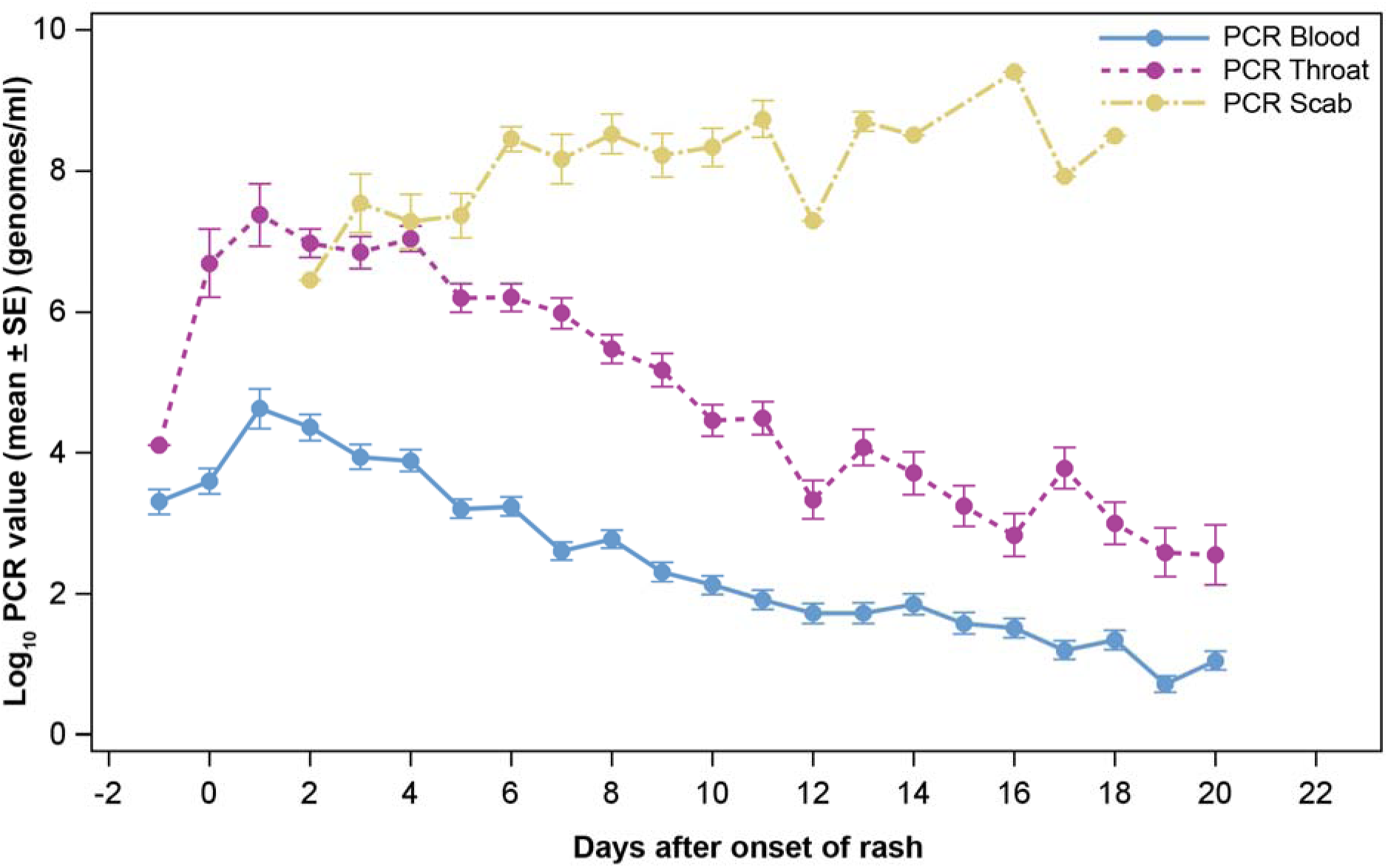
Change in PCR count from blood, throat and scab with time. Mean of PCR count from blood, throat and scab (Log_10_) with standard error bars (Unit: genomes/mL) over time.

There was no difference in the level and decline over time for males and females for blood (p = 0.9079) or pharynx (p = 0.5208) PCR copy numbers. The Wilcoxon-Mann-Whitney test showed no significant differences in maximum PCR blood or maximum PCR throat swabs results (p = 0.4505 and 0.8778, respectively). Scabs contain significant quantities of MPXV positive DNA until and including when they fall off. The concentration of MPXV in the scab is several times the number of genomes in blood and throat. Viral infectivity in specimens was not determined.

### Secondary household cases of Monkeypox

For this analysis, patients who developed skin rash 14 days or longer after the first household case of monkeypox were labeled secondary cases. Of the 216 patients in the study, 105 had family relationships. Twelve of the 44 families (27.27%) or 18 family members in this study had secondary infections by this definition. The rate of secondary infection patients compared to the total number of patients in the study is 8.34% (18/216). Patients diagnosed as

secondary cases had a lower mean total lesion count (386 vs 618) that peaked at day 4-5 and healed at a faster rate than primary cases. Primary cases had a higher total lesion count (8619 vs 2448) that peaked at day 9-10 and healed more slowly compared to secondary cases (p = 0.1373). Classic primary and secondary cases are presented below.

- A hunter presented to the hospital with primary MPX and a fever of 7 days and pox lesions present for 5 days with an admission lesion count of 427 that peaked on lesion day 10 with 751 lesions. On admission the patient had severe lymphadenopathy with blood MPX PCR viral load 2.1 X 10^5^ genomes/ml, the maximum value seen in this patient that slowly decreased to 1.5 X 10^3^ by lesion day (LD) 19. The patient was discharged after 34 days of hospitalization, much longer than normal.
- A household member, presented with secondary MPX with enlarged cervical lymph nodes (10 mm), positive blood MPX PCR viral load (4.8 10^3^ genomes/mL) and throat swab (1.4 X 10 genomes). Lesions were not present on admission and first appeared on the 3^rd^ hospital day (HD) LD 0 with 3 lesions that reached a maximum on HD 6 (LD 4) with 21 lesions and blood genomes reached a maximum on HD 5 with 1.3 X 10^5^ genomes/ml that slowly decreased to 3.9 X 10^5^ on discharge, HD 18. These 2 cases will be the subject of a more detailed report to be published separately.

### Pregnancies and fetal outcomes

The course and outcome of four cases of maternal MPXV infection were described in a previous publication (26). One of the four cases survived its mother’s monkeypox infection. Two pregnancies resulted in spontaneous abortions. The fourth suffered intrauterine death. There was PCR and histopathologic evidence of a high monkeypox viral load in this fetus (26) (Figure 9 A-C--photographs are cropped-consult authors for full de-identifiable photographs)

**Figure 9.**
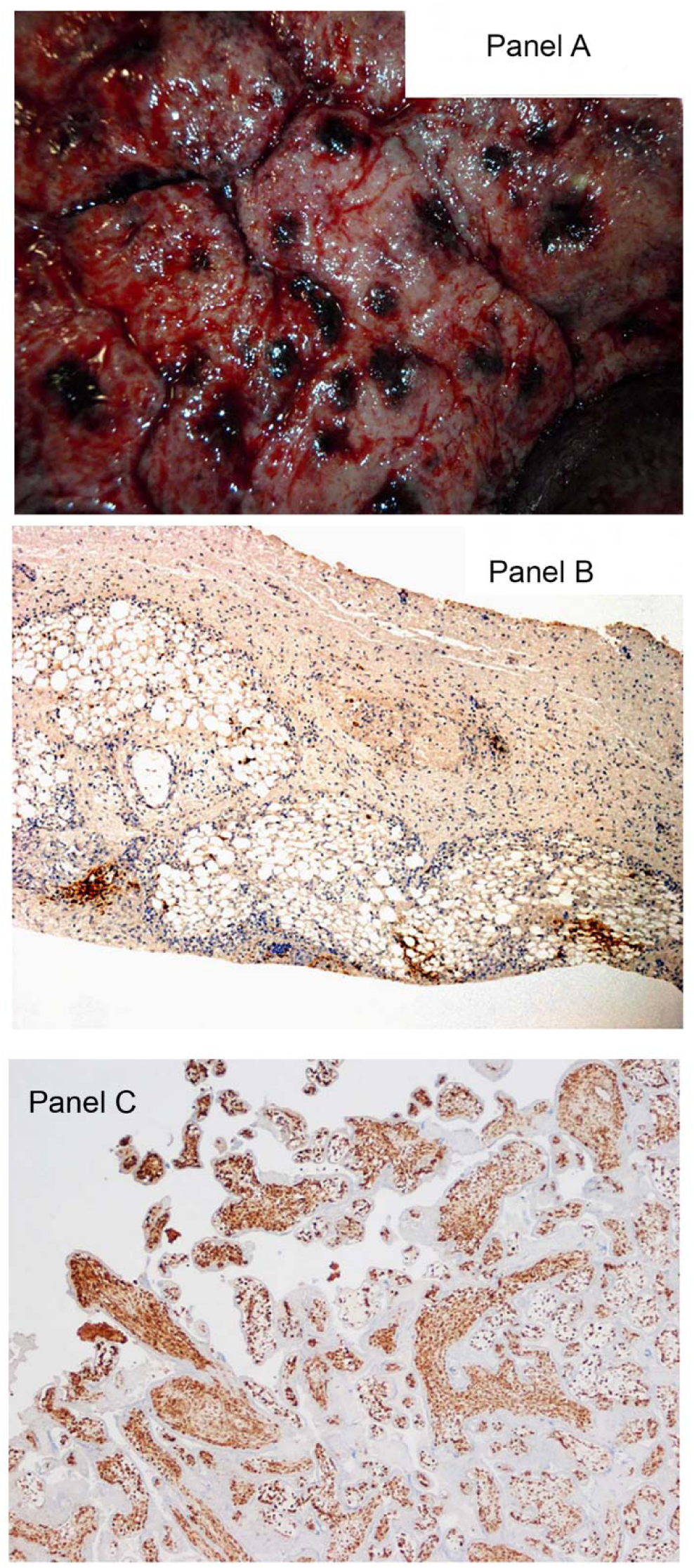
The evidence of Monkeypox infection on Fetus and Placenta. (A) Maternal surface hemorrhages of placenta (B) There is diffuse epithelial positivity on placenta. 4X objective. (C) There is multifocal positivity in the fetal skin. 10X objectives. Panels B & C are Vaccinia Immunohistochemical examination of formalin fixed thin sections stained with H&E.

### Monkeypox virus infection death

Three deaths occurred among the 216 patients in this cohort for which a detailed accounting will be presented in a future publication. Two deaths occurred in the 5-11 year old group, one in the <5 year old group and none in the ≥12 year old age group. The 3 deaths are described briefly below:

Fatal Case #1: A gravely ill pediatric patient had 2091 lesions (VES 578, PUS 1375, OMB 37) on admission at 6^th^ day of fever and 5^th^ day of rash. The patient’s viral load by PCR was 2.4 X 10^6^ viral genomes/mL blood on admission. By day 2 lesions had increased to 2447 and fever had resolved. The patient developed respiratory distress and agonal breathing by 1700 and died at 2200 with elevated AST of 185 (2) and ALP of 304. Figure 9 A-C shows lesions on maternal surface of placenta and microscopic findings of placenta and infant skin lesions.

Fatal Case #2: A pediatric patient was admitted with complaints of fever for 3 days with onset of pustular lesions. More than 1,000 vesicular and pustular lesions, along with a few umbilicated lesions were observed. The clinical assessment was a child with severe early-stage monkeypox, dehydration, ketoacidosis and proteinuria. MPX viral genomes in blood were initially 10 genomes/mL and remained close to that value until death (this case will be reported in detail in a later manuscript).

Fatal Case #3: A third pediatric patient, part of a family cluster of 4 patients was admitted on the 4^th^ day of fever and 3^rd^ day after lesions developed. The patient had a very high viral load (2 X 10^6^) on admission, 601 lesions and died the next day with very high liver transaminase values (AST = 865).

Death occurred in one pediatric patient from apparent respiratory illness 9 days after discharge from hospital with monkeypox at which time the patient was disease free. The death occurred at a location remote to the study site and investigators only became aware of the child’s death when the family did not return for the post-hospitalization follow-up visit. The cause of death cannot be confirmed as MPX. Although, orthopoxvirus induced immunosuppression is suspected as having contributed to the patient’s death by investigators. This case will be discussed in detail in a later publication.

### Comparisons between patients with fatal disease and survivors

All patients who succumbed to MPX disease complained of malaise, sore throat and anorexia compared to 50-84% of survivors but this difference did not reach statistical significance. Except for the number of MPX lesions, there were no S&S that distinguished between those who survived and those who died of MPX infection. Maximum lesion count was significantly different between patients who survived and those who succumbed to MPX (p = 0.0025). Fatal cases had a significantly higher lesion counts (GM = 2,294, 95%CI: 79-66,842) than surviving patients (GM = 195, 95% CI: 162-235). There was a significant difference in maximum blood PCR genome levels between patients who succumbed and those who survived MPXV infection (p=0.0072). Patients who died had significantly higher maximum PCR blood genome levels (geometric mean (GM) = 9,204,937, 95%CI: 2.1x10^4^-4.01x10^9^) than surviving patients (GM = 22,971, 95%CI: 1.4x10^4^-3.8x10^4^). The mean values for ALT and AST for patients who died or survived were: 90 vs 26 U/L (p = 0.0224) and 415 vs 48 U/L (p =0.0002) (p-values, adjusted), respectively.

### Association analyses

Figure 10 displays the Odds Ratio (OR) and p-values related to the association analyses. The OR is one way to present the strength of association between risk factors/exposures and outcomes. It represents the odds that an outcome will occur in the presence of a risk factor or exposure, compared to the odds of the outcome in the absence of a risk factor or exposure. If the 95% confidence interval for an OR includes 1, it means the results are not statistically significant. Figure 10 shows the significant associations found in this analysis. Level of consciousness (lethargy and/or stupor) has the highest ORs relative to its presence and disease severity (ORs up to 24.34; P<0.0001); followed by activity level (Incapacity) with ORs up to 6.32. Details of certain subcategories are shown as additional Forest Plots in Supplemental materials (Figures S1A-D).

**Figure 10.**
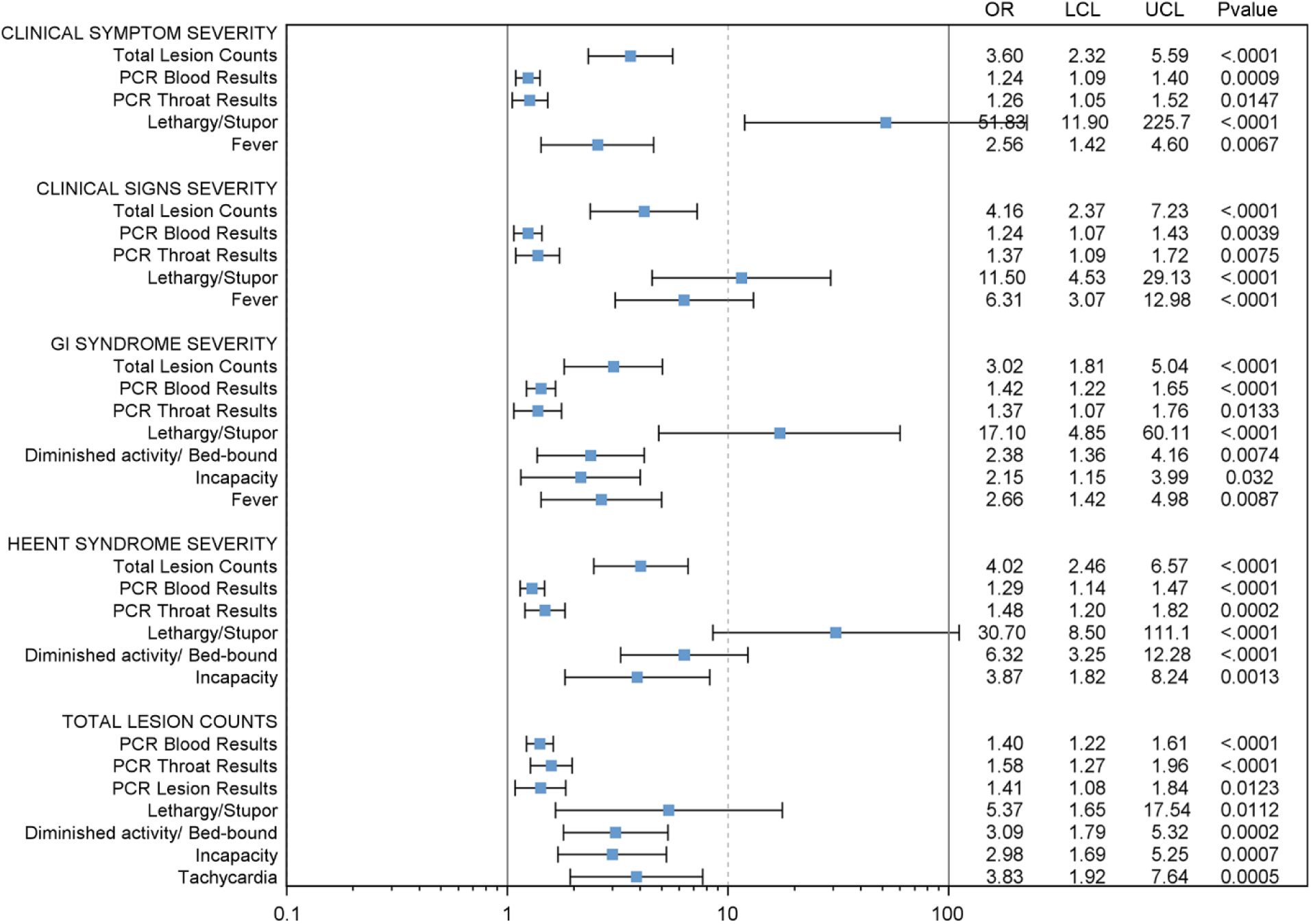
Significant associations found in the analysis. Forest Plot showing statistically significant associations. Odd ratio (OR), upper 95% confidence interval (UCL), and lower 95% confidence interval (LCL) were calculated using separate generalized estimating equations (GEE) with cumulative logit models, day after rash onset was adjusted as covariate. If a 95% confidence interval for an OR is >1, the odds of a given outcome are increased in the presence of a risk factor or exposure. If the 95% confidence interval for an OR is <1, it means the odds of a given outcome are decreased in the presence of a risk factor or exposure. Adjusted *p* value were calculated using stepdown Bonferroni correction if needed. Only significant results are showed here (*p* value ≤ 0.05).

## Discussion

The most complete early description of human MPX infection comes from Bremen’s description of 47 cases between 1970-1979 in West and Central Africa and Ježek’s 338 cases between 1981-1986 from the two WHO MPX study sites in Zaire (opened after the eradication of smallpox, reported in 1980) (27, 28). The epidemiologists described human MPX as closely resembling, discrete ordinary-type smallpox or occasionally modified type smallpox. There have been reports of several cases of morphologically atypical lesions in human MPX patients including a previously reported case who was a stillborn infant from this study. No cases of human MPX have yet been reported that resemble either confluent or hemorrhagic smallpox either in the appearance of the exanthem or the rapidly fatal clinical course. The results of this investigation reinforce prior observations of the severity and systemic nature of this infection, while also highlighting opportunities to ameliorate clinical care and patient management.

The most characteristic and distinguishing characteristic of MPX from smallpox has been the appearance of larger numbers of cases of MPX with painful, tender lymphadenopathy most frequently noted in the cervical and submandibular regions, but also occurring as inguinal adenopathy or generalized distribution, 98.6% of patients had lymphadenopathy. Our study validates previous observations regarding lymphadenopathy occurrence in MPX as shown in Figure 6. Enlarged lymph nodes may compress the airway leading to respiratory compromise. Parental steroids may be indicated in these situations. Patients with multiple oropharyngeal lesions may refuse to eat or drink as well and may become dehydrated. Intensive nursing care and hydration would benefit these patients.

The PCR viral load from the throat were about 2000 genomes/mL more than blood supporting the idea of swabbing the throat especially when MPXV infection is suspected but blood PCR values are low or absent. As with smallpox, the scabs of MPV are highly concentrated with virus even when it falls off. Maximum PCR values are known to correlate with outcome in the lethal cynomolgus primate model including significant reductions with successfully antiviral drug treatment (7, 29, 30). Maximum values occur early in the disease course which can be accurately determined in controlled animal studies, however patients entered the study after rash onset at a point at which the peak viremia had passed resulting in the study failing to capture viral load peak levels for many patients. Thus, being admitted several days after rash onset detracted from our ability to statistically validate maximum PCR with certain disease indicators (consistent with early admissions having higher maximum PCR values). Early disease seen in one patient supports the idea that maximum PCR values (a measure of viral load) occur very early in the disease course. Patients admitted several days after lesion onset are already past their maximum PCR value which explains why patients admitted closer to the onset of their rash tended to have higher maximum detected PCR values. For this study, the true maximum viral load was not determinable due to late admission of patients with MPX. On the other hand, for most patients with non-fatal disease, maximum <u>lesion</u> counts determined later in the course of the illness appeared to correlate with non-fatal disease severity more clearly than observed maximal <u>viral</u> load.

Exposure to 2 or more wild animals was noted in 94.9% of patients [Table 1]. Bremen reported the rate of interhuman transmission to susceptible household members as being 7.5% in his series (27). In this report, we found the rate of secondary household transmission to be 8.3%. Three-quarters (72.7% of households with an index case of monkeypox did not suffer secondary infections in our study.

Human MPXV infections result in a wide spectrum of illness ranging from mild to severe and even death in this hospitalized population. Wide variation was observed in lesion counts. The classic MPXV rash or lesion was observed in 99. 5% of patients. The single patient with MPX but without lesions was determined to have been vaccinated against smallpox decades earlier. Because smallpox had largely disappeared from the developed world by the beginning of the 20^th^ Century, little is known or understood regarding extent of organ systems effected directly by VARV, or how isolated impairment of for instance liver or renal function may have contributed either to the morbidity of the illness or to also the mortality from infection. This study used routine hospital laboratory testing to evaluate the occurrence of organ system dysfunction and compare to clinical parameters of disease morbidity. Higher elevations of ALT and AST did correlate with mortality, but the enzyme elevations were consistent only with moderate acute liver injury and other parameters did not correlate with hepatic failure. While dehydration was likely a major contributor to mortality, infection induced renal failure did not appear to play a role in events leading to a patient’s demise. Death in MPXV infections does not appear to be caused by progressive multisystem failure based upon these observations and evidence of multisystem failure is only seen as a pre-terminal event.

The predictors of poor prognosis included elevated AST and ALT; patients with these findings should guide medical care providers toward intensive supportive care. Patients with multiple oropharyngeal lesions and large cervical and other neck region lymphadenopathy who are at greater risk of respiratory compromised, will not want to eat or drink as well and may become dehydrated. Because motorized transportation is not available, many patients arrived at the hospital, dehydrated, malnourished, anorexic, and fatigued, adding to the physiologic stress of infection.

There were very few deaths in this case series, consequently we were not able to draw firm conclusions regarding symptoms or physical findings that correlate with fatal infections. The day of onset of symptoms prior to hospitalization was not collected, therefore, the sequence of symptoms cannot be determined by this dataset, nor can symptoms duration be precisely determined, although we can capture duration from the day of hospitalization. Likewise, symptom severity was not collected. We used the number of symptoms as a surrogate, based upon a symptoms vs signs matrix. We graded the illness using cumulative totals of types of symptoms and numbers of individually identifiable signs. Using this arbitrary grading system, we classified patients as level 1, level 2, level 3 and level 4 (fatal) (Table 3A and 3B) and present data reflective of the illness severity categories. Patients with higher total number of S&S should be considered for hospitalization even if resources are limited. Patients who are confused/disoriented/lethargic or stuporous had the highest Odds Ratios (OR’s range 5.7-30.74) and should be provided immediate and intensive care. Frequently occurring symptoms included sore throat anorexia, cough, fever, chills, nasal discharge and congestion, dysphagia, mouth/throat lesions, headache, abdominal pain, sweats conjunctiva lesions, shortness of breath. Frequently occurring physical findings were diminished levels of physical activity, nonspecific rash, mouth/throat lesions, fever, abnormal lung sounds, hepatomegaly/splenomegaly, lethargy/stupor, dehydration, confusion and or disorientation. On a case-by-case basis, patients without these symptoms and signs could be considered for triage to home treatment, providing hygiene, hydration and nutritional support are available within the family unit. There is still much clinical work needed to refine and validate a grading system to provide decision support regarding disposition and treatment of individual patients. Based on the severity of disease, in young children, any grading system for triage, should take the age of the patient into account as one of the major factors in determining the need and benefit of hospital based care.

The case fatality rate of 1.4% (3/216) is markedly lower in this hospitalized cohort than has occurred historically. The death of a pediatric patient occurred 9 days after being discharged free from monkeypox disease. While being carried by its parents, the child developed an acute respiratory infection during the long journey home and died 3 days after arrival at their village. A case-fatality rate of 9.8% (33/338) was noted during a WHO sponsored study in Zaire during 1981-1986 (26). A later study sponsored by the WHO during 1996-7 had a case-fatality rate of 3.7% (27).

Fetal demise, the topic of a previous publication, occurred in 3 of 4 (75%) pregnancies due to maternal MPXV infection [26]. In spite of confirmation of the relationship between pregnancy and severe and fatal infection maternal MPXV infection was not always fatal to the mother or the fetus.

Co-infections were frequent and included, chickenpox, malaria, microfilariasis, giardiasis, ankylostomiasis, strongyloidiasis, filariasis species, and hookworm. Skin infections were often present, including infected wounds. Several complications were noted such as, caseation of the eye, keratitis, etc. A later publication will describe the MPXV-VZV co-infection cohort that occurred during the course of this study.

A vast amount of data were collected during the conduct of this observational study much of which cannot be adequately presented in this publication. Future publications will include analysis of deaths associated with MPXV infections during this study, coinfections such as chickenpox, complications observed among patients during this study, detail analyses of the antibody data including peptide data, cytokine data, etc.

Recent efforts have facilitated FDA licensure of Jynneos, a non-replicating, third generation vaccinia vaccine, for the prevention of smallpox and monkeypox (31, 32). Studies are underway to examine the efficacy of this vaccine in prevention of human MPX (33). Additionally, Tecovirimat (TPOXX, ST-246) and Brincidofovir were recently licensed by FDA for the treatment of poxviral infections (34). Early treatment to prevent serious MPXV infection should be investigated. As with other infectious diseases and intoxications, treatment before or proximal to the onset of symptoms have the best outcomes.

## Data Availability

All relevant data are within the manuscript and its supporting Information files. Data are also available from the Office of Human Research Oversight, USAMRIID for researchers who meet the criteria for access to confidential data.

## Acknowledgments

We are most grateful to Dr. Therese Riu-Rovira for serving as medical monitor for the study. Dr. Riu-Rovira is one of the two founding Missionaries of Christ Jesus, a Spanish Order of Catholic Sisters, who have been managing the hospital and cases of human monkeypox disease for over 30 years. We thank Diana Fisher for her expert and professional statistical recommendations and support. Our sincerely thanks to Staff Sergeant Keith Kittle and Staff Sergeant Jesse Kaplan for infrastructure buildout and maintenance.

## Kole Human Monkeypox Study Group

Jules Alonga, Guy Bilulu, Diana Blau, Jillybeth Burgado, Russ Byrne, Gary Carter, Matthew S. Chambers, Jennifer Chapman, Didace Demba, Francis Devlin, Pierre Ewala, Kathleen Farr, Robert Fisher, Steven Gire, Tim Haley, Lisa Hensley, Paul Hollier, Monique Jesionowski, Jesse Kaplan, Keith Kittle, James Koterski, David Kulesh, Keith Iams, Donnat Mbale, Paul Melstrom, Mitch Meyers, Eric Mucker, Gaston Mwema, Emile Okitolonda, Eddy Ortega, Kerry Patterson, Nishi Patel, Sister Marisa Ramos, Sister Therese Ruis, Shirley Roach, Thomas Robinson, Kate Rubins, John Schaber, Karen Sellers-Myers, David Sugerman, Max Teehee, James Wadding, Sean Wilkes, Mark Withers.

## Disclaimer

Opinions, interpretations, conclusions, and recommendations are those of the authors and are not necessarily endorsed by DOD or HHS (CDC). Research on human subjects was conducted in compliance with U.S. Department of Defense, federal, and state statutes and regulations relating to the protection of human subjects and adheres to principles identified in the Belmont Report (1979).

---

**Supplemental Figure S1A.**
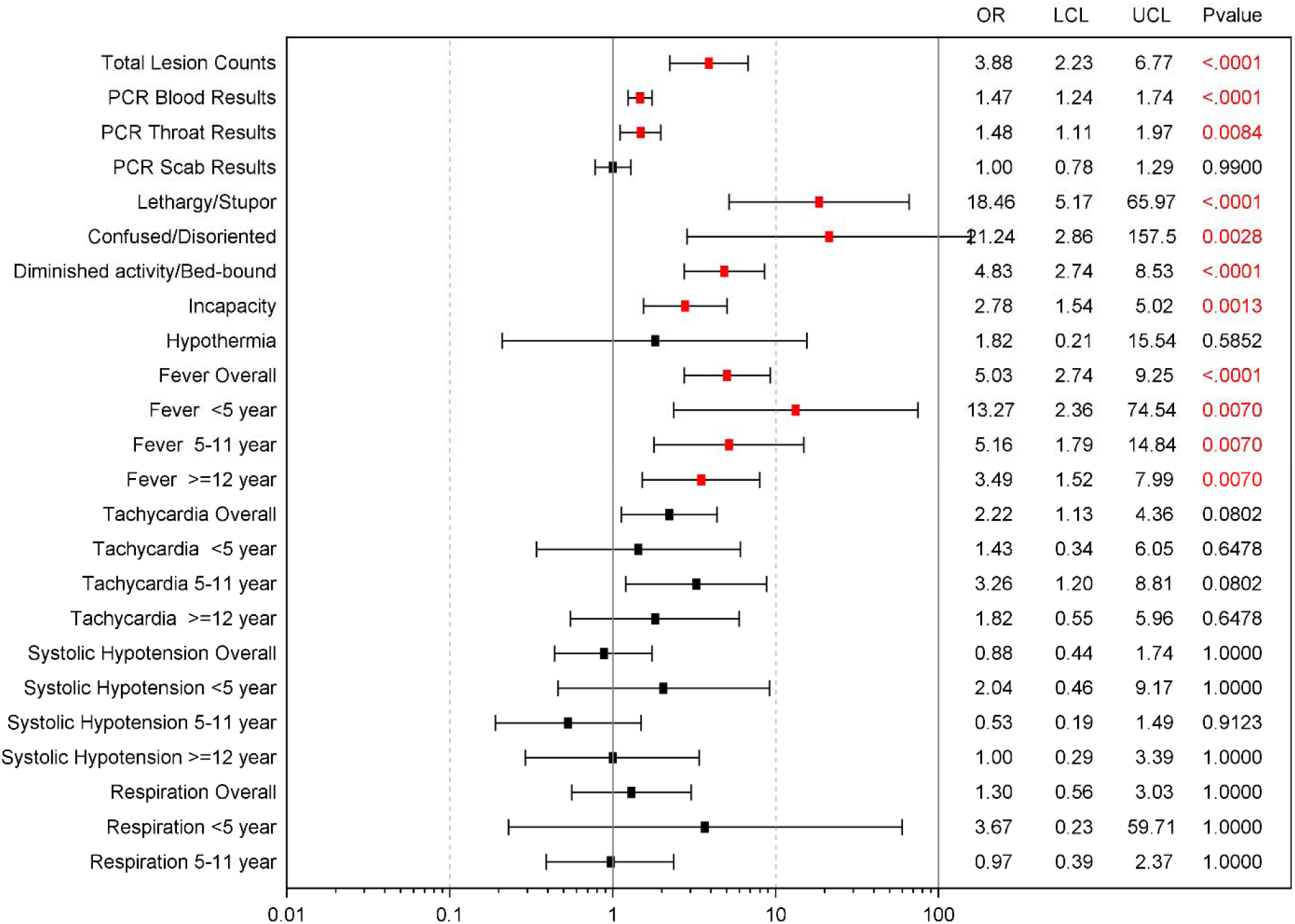
Associations between monkeypox illness severity categories and other variables. Forest Plot showing associations between monkeypox illness severity categories and other variables on the admission day. OR, UCL and LCL were calculated by GEE with cumulative logit models, day after rash onset were adjusted as covariate and *p* value were adjusted by stepdown Bonferroni correction.

**Supplementary Figure S1B.**
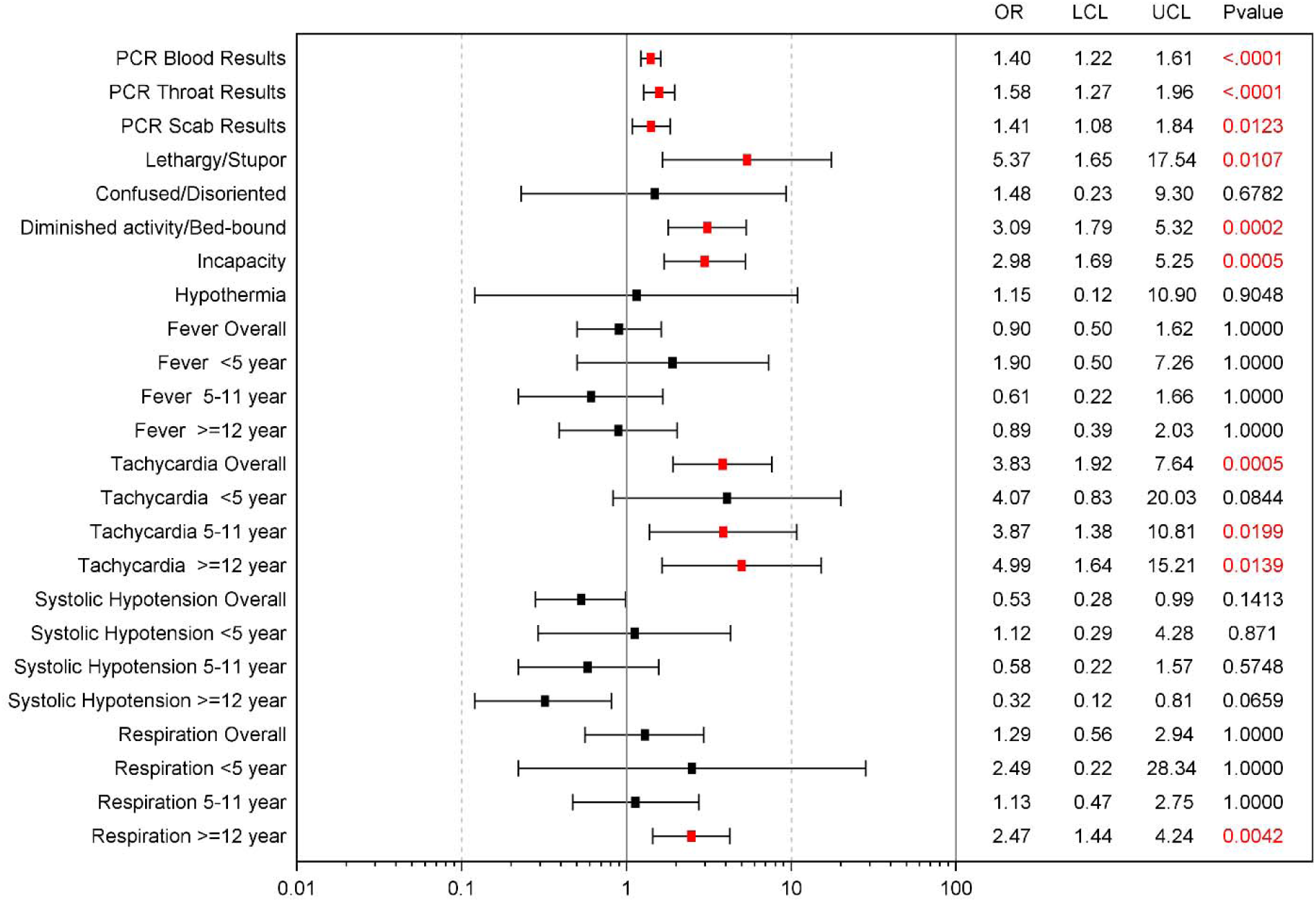
Associations between total lesion count severity with other variables. Forest Plot showing associations between total lesion severity and other variables on the admission day. OR, UCL, LCL were calculated using GEE with cumulative logit models, day after rash onset and age group were adjusted as covariate and *p* value were adjusted by stepdown Bonferroni correction.

**Supplementary Figure S1C.**
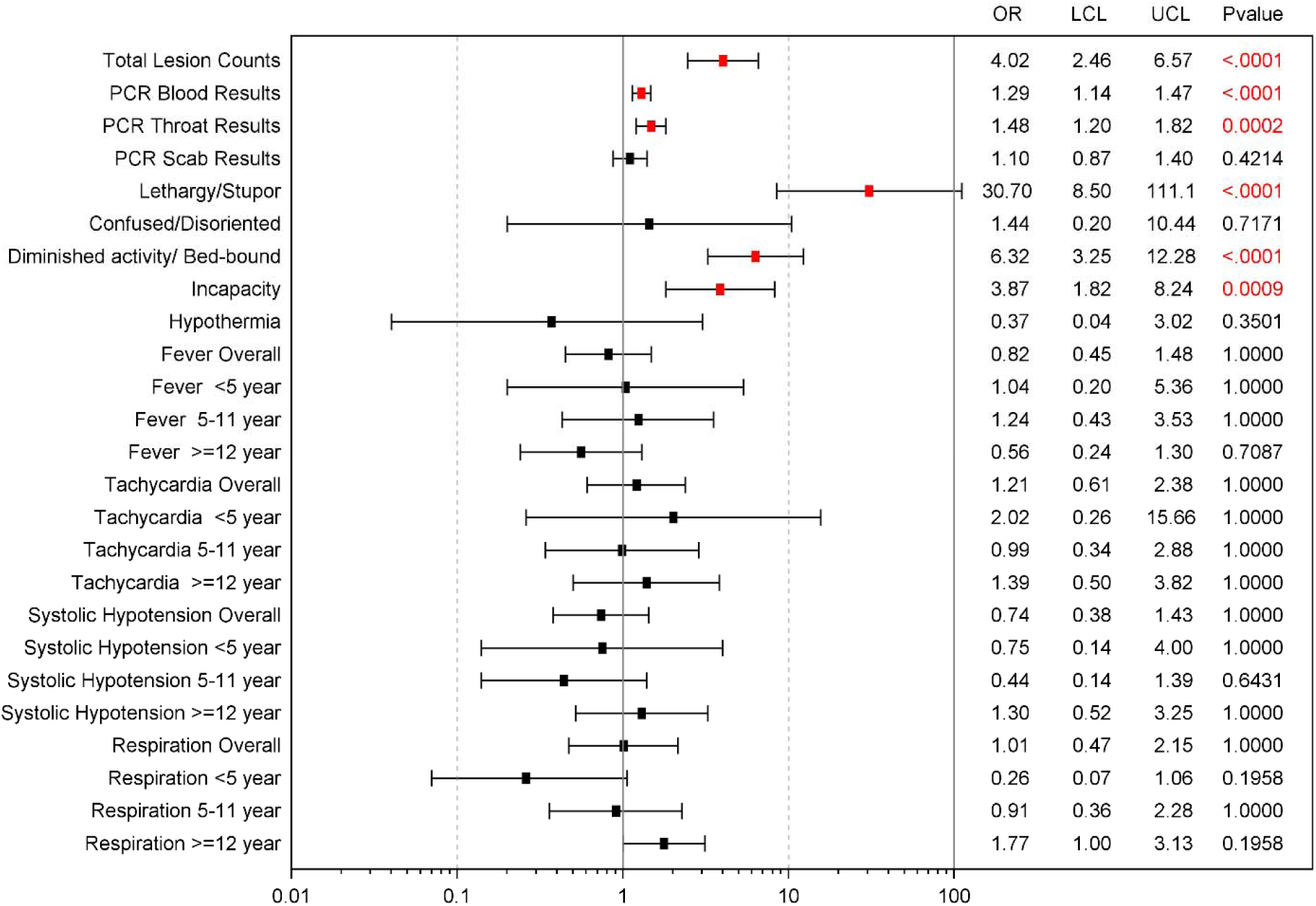
Associations between HEENT syndrome severity with other variables. HEENT included following clinical symptoms or signs: Visual changes, Eye pain/discharge, Ear pain, Nasal discharge/congestion, Dysphagia, Sore throat, Conjunctive and other eye lesion, Nasal discharge/congestion/ rhinorrhea/nasal lesion, Mouth/throat lesions. The HEENT severity scores are based on the number of abnormal HEENT: Grade 0 (0), Grade 1 (1-2), Grade 2 (3-5), Grade 3 (6-7), Grade 4 (8-9).

**Supplementary Figure S1D.**
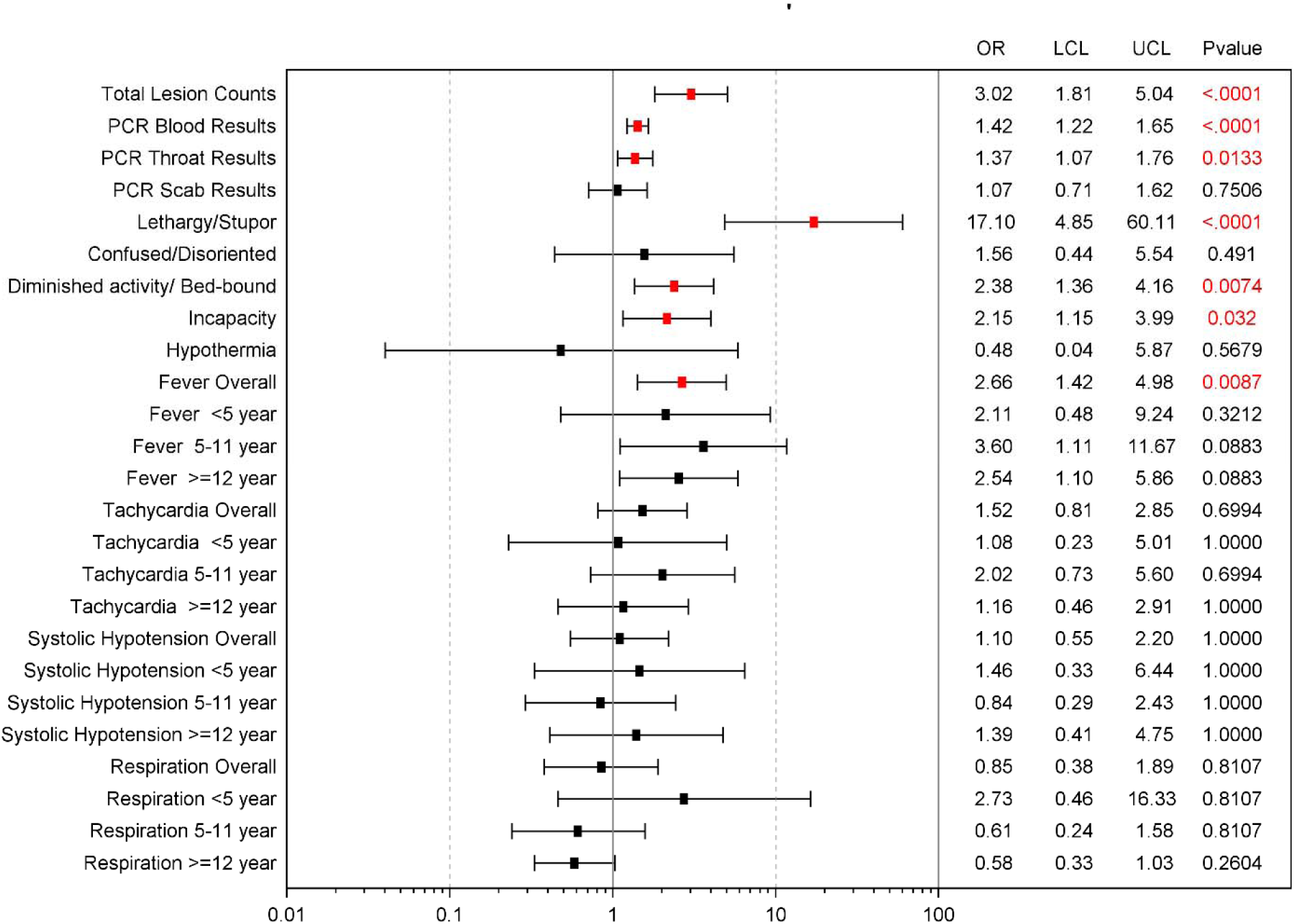
Associations between GI syndrome severity with other variables. GI included following clinical symptoms or signs: Anorexia, Vomiting, Abdominal pain, Dysphagia, Diarrhea, Hepatomegaly, splenomegaly or both, Abdominal tenderness. The GI severity scores are based on the number of abnormal HEENT: Grade 0 (0), Grade 1 (1-2), Grade 2 (3-4), Grade 3 (5-8).

**Supplemental Table S1.** Defining disease severity categories.

| Symptom<br>Sign | 0 | 1 | 2 | 3 | 4 | 5 | Total |
| --- | --- | --- | --- | --- | --- | --- | --- |
| 4 | 0 | 0 | 0 | 1 | 4 | 1 | 6 |
| 3 | 0 | 2 | 1 | 21 | 10 | 3 | 37 |
| 2 | 1 | 16 | 35 | 56 | 7 | 1 | 116 |
| 1 | 5 | 22 | 19 | 7 | 0 | 0 | 53 |
| 0 | 0 | 1 | 0 | 0 | 0 | 0 | 1 |
| Total | 6 | 41 | 55 | 85 | 21 | 5 | 213 |

**Supplementary Table S2.** Descriptive tables of Clinical laboratory values in patients with acute monkeypox infection on admission day.

| Laboratory Test (Unit) | N | Mean(SD) | Median | Min | Max | Normal Range |  |
| --- | --- | --- | --- | --- | --- | --- | --- |
|  |  |  |  |  |  | Male | Female |
| ALB (G/DL) | 206 | 3 (0.5) | 2.75 | 1.3 | 7.7 | 3.3 - 5.5 | 3.3 - 5.5 |
| ALP (U/L) | 202 | 123 (48.2) | 112 | 0.5 | 304 | 53 - 128 | 42 - 141 |
| ALT (U/L) | 206 | 27 (22.6) | 22 | 2.5 | 201 | 10 - 47 | 10 - 47 |
| AMY (U/L) | 205 | 78 (86.8) | 61 | 2.5 | 1061 | 14 - 97 | 14 - 97 |
| AST (U/L) | 205 | 54 (77.4) | 39 | 9 | 865 | 11 - 38 | 11 - 38 |
| BUN (MG/DL) | 187 | 8 (4.7) | 7 | 0.5 | 32 | 7 - 22 | 7 - 22 |
| CA (MG/DL) | 205 | 9 (0.7) | 8.9 | 2.7 | 10.2 | 8 - 10.3 | 8 - 10.3 |
| CRE (MG/DL) | 202 | 1 (0.4) | 0.6 | 0.1 | 5.5 | 0.6 - 1.2 | 0.6 - 1.2 |
| GGT (U/L) | 206 | 47 (65.0) | 30 | 2.5 | 703 | 5 - 65 | 5 - 65 |
| GLU (MG/DL) | 206 | 94 (21.7) | 92 | 41 | 209 | 73 - 118 | 73 - 118 |
| TBIL (MG/DL) | 205 | 1 (0.3) | 0.6 | 0.3 | 2.4 | 0.2 - 1.6 | 0.2 - 1.6 |
| TP (G/DL) | 206 | 8 (0.9) | 7.8 | 3.2 | 13 | 6.4 - 8.1 | 6.4 - 8.1 |

